# Systems immunology reveals the molecular mechanisms of heterogeneous influenza vaccine response in the elderly

**DOI:** 10.1101/2023.07.10.23292445

**Authors:** Saumya Kumar, Martijn Zoodsma, Nhan Nguyen, Rodrigo Pedroso, Stephanie Trittel, Peggy Riese, Javier Botey-Bataller, Liang Zhou, Ahmed Alaswad, Haroon Arshad, Mihai G. Netea, Cheng-Jian Xu, Frank Pessler, Carlos A. Guzmán, Luis Graca, Yang Li

**Author notes:** Correspondence (Y.L.). These authors contributed equally (Saumya Kumar, Martijn Zoodsma). LeaSd contact (Yang Li).

## Abstract

Vaccination-induced protection against influenza is greatly diminished and increasingly heterogeneous with age. We investigated longitudinally (up to five timepoints) a cohort of 234 elderly influenza vaccinees across two independent seasons including up to six modalities (multi-omics and immunological parameters). System-level analyses revealed responders exhibited time-dependent changes attributed to a productive vaccine response across all omics layers whereas non-responders did not follow such dynamics, suggestive of systemic dysregulation. Through multi-omics integration, we identified key metabolites and proteins and their likely role in immune response to vaccination. High pre-vaccination IL-15 concentrations negatively associated with antibody production, further supported by experimental validation in mice revealing an IL-15-driven NK-cell axis with a suppressing role on antibody production. Finally, we propose certain long-chain fatty acids as modulators of persistent inflammation in non-responders. Our findings highlight the potential for stratification of vaccinees and open avenues for possible pharmacological interventions to enhance vaccine responses.

**HIGHLIGHTS:**

1. Pre-vaccination pro-inflammatory status impedes vaccine responsiveness in the elderly.
2. Multi-omics integration reveals key molecules involved in vaccine response.
3. High pre-vaccination concentrations of IL-15 suppresses vaccine response through NK cell activation.
4. Certain long-chain fatty acids may act as modulators against chronic inflammation and are potential targets to improve vaccine response.

## INTRODUCTION

Influenza epidemics are major public health threats, with more than 400,000 fatalities estimated worldwide per year, mostly among the elderly^1^. Inactivated subunit influenza vaccination has proven to be the best preventive and cost-effective approach for inducing protective immune memory^2, 3^. However, many factors, particularly old age, have been strongly associated with a reduced protection despite vaccination^4, 5^. Consequently, the elderly population is more vulnerable to influenza infection and influenza-associated mortality^1^.

The inefficiency in generating protective immunity in the elderly has been attributed to the ageing of the immune system, which is often characterised by lingering low-grade inflammation and immunosenescence^6^. Our recent studies also showed that non-responders are specifically characterized by multiple suppressive immune mechanisms affecting regulatory T and B cells^7^. Earlier studies have also shown higher proportions of inflammatory monocytes and cytotoxic NK cells in the elderly compared to the younger population, along with lower B cell responses^8^ that were attributed to intrinsic defects in T and B cells. Additionally, the elderly accumulate pro-inflammatory B cells that are unable to respond to influenza vaccination^9, 10^. At the molecular level, lower AP-1 activity along with enhanced antiviral and interferon signalling in myeloid cells has been associated with a productive vaccination response^11^. Higher AP-1 activity induces activation of pro-inflammatory cytokines, which may contribute towards a higher inflammatory status^11, 12^. However, in the younger population (< 50 years), pre-vaccination pro-inflammatory status driven through NF-kB signalling has been associated with high antibody response^13^. In addition to cytokines, metabolites may also serve as important regulators of immune responses^14, 15^. Differences in metabolite profiles between young and elderly individuals have been reported, along with differential metabolic regulation of the immune response to vaccination^16^. Post-vaccination, young high responders showed reduced concentrations of polyunsaturated fatty acids (PUFAs), whereas cholesteryl esters accumulated more in elderly high responders^16^. Purine metabolism and glycine, serine and threonine metabolism have also been associated with the vaccine response^16^. Together, these studies have indicated complex and age-dependent metabolic regulation of the antibody response to vaccination.

Here, we used a systems immunology approach to examine the differential responsiveness to influenza vaccination within a previously well-established elderly cohort spanning two independent influenza seasons^7, 17–19^. The transcriptional signatures of the highest and lowest responders allowed us to identify a chronic inflammatory signature that is distinct from the inflammatory signature supporting the vaccine response. We also show distinct changes in the plasma proteome and metabolome abundance upon vaccination in high and low responders. Furthermore, we identified pre-vaccination protein biomarkers associated with poor vaccine response. Additionally, we identified pre-vaccination metabolites associated with vaccine response, which may serve as potential modulators of the chronic inflammation in the elderly. Taken together, these findings illustrate that an increased pro-inflammatory profile is detrimental to influenza vaccine response in the elderly population. Results from this study provide insight into the immunological processes driving differential responsiveness to vaccination, and will open novel opportunities for improved vaccine design as well as modulation of the immune system to enhance the vaccine response in the elderly.

## RESULTS

### Demographic characteristics and their association to vaccine response in the elderly

We collected whole blood and plasma samples from 234 trivalent inactivated influenza vaccine (TIV)-vaccinees aged 65 and above in Hannover, Germany across two influenza seasons and generated a multimodal dataset covering whole blood transcriptome, plasma proteome, plasma/serum metabolome, and serology from up to five time points (pre-vaccination, days one-three, six-seven, 21-35 and 60-70 post-vaccination). By doing this, different stages of the vaccination response were covered (Figure 1A). Examining the serological data, we observe heterogeneity across the elderly donors, with the majority of donors showing sufficient antibody responses expected to confer protection. Donors were evaluated based on their antibody fold change (day 35 vs pre-vaccination) against the three strains included in the TIV vaccine^7^, and categorised as Triple Responders (TR, >4-fold rise in antibody titres against all three strains), Non Responders (NR, <4-fold rise in titres against all three strains), or Other (>4-fold rise in titres against one or two of the vaccine strains; Figure 1B). As we have access to data from two independent seasons, we used the larger cohort (season 2015/2016, n=200) as a discovery cohort, and the smaller cohort to replicate our findings (season 2014/2015, n=34).

**Figure 1.**
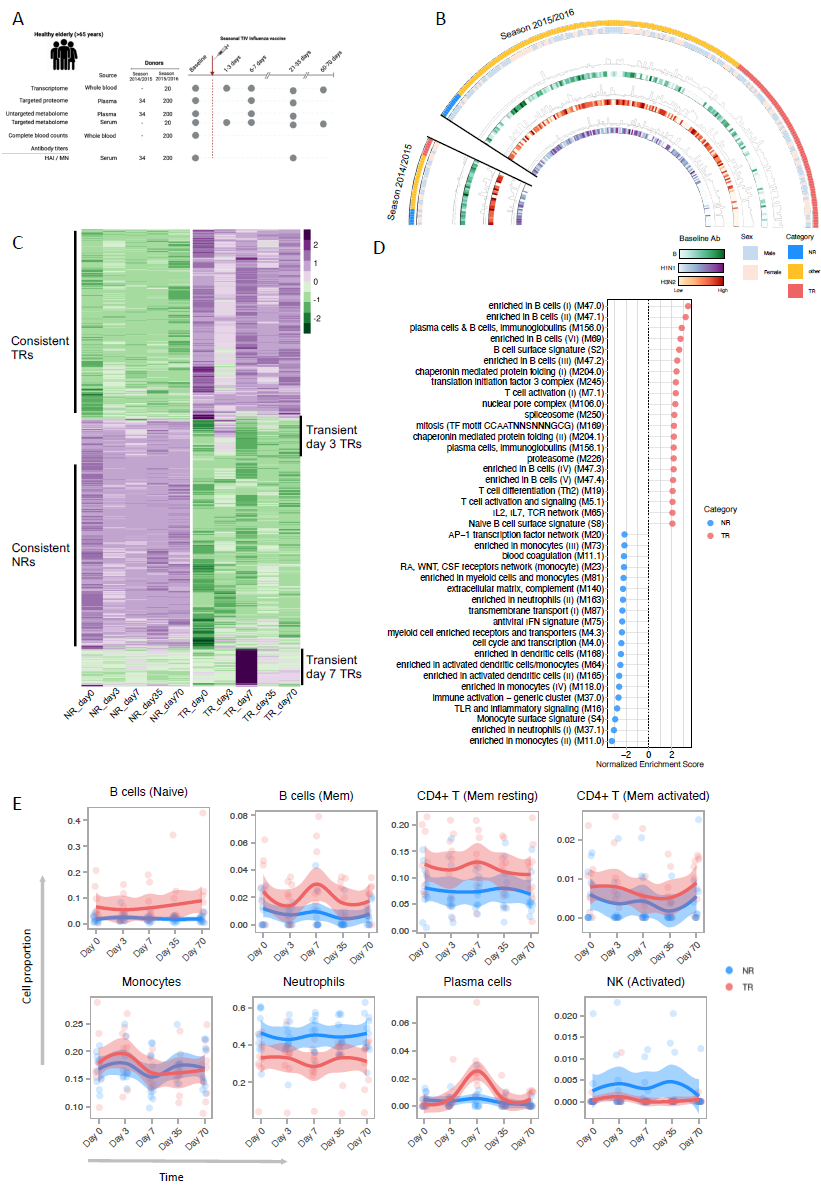
Influenza vaccination cohort overview and transcriptome of highest and lowest responders. (**A).** Overview of the different omics datasets generated in this study. We generated multi-modal datasets covering the transcriptome, proteome and metabolome response to influenza vaccination from up to 234 individuals for different timepoints spanning both pre and post-vaccination, covering two independent seasons of influenza vaccination. The serological data is adapted from earlier publications on the same cohort^7, 17^ (**B).** Circos plot showing the serological response to trivalent inactivated influenza vaccine (TIV). Each heatmap depicts pre-vaccination antibody titres against the particular strains included in the vaccine (H3N2, H1N1, B), whereas the corresponding histograms show the log2 fold-change in antibody titres against the strains upon vaccination. Donors were classified as TR, NR or Other based on their serological response (outer tiles). (**C)** Heatmap showing mean gene expression levels across 10 TRs and 10 NRs for each time point. Each row is a gene that is significantly differentially expressed between TRs and NRs, with purple indicating higher gene expression levels and green indicating lower gene expression levels (adj. p-value < 0.01). (**D)** Top 40 BTMs listed based on gene set enrichment analysis (GSEA) of genes that are differentially expressed between TRs and NRs. Positive (red) normalised enrichment scores (NES) correspond to BTMs upregulated in TRs, whereas negative NES corresponds to BTMs upregulated in NRs. (**E)** Line plots depicting immune cell population proportions for all timepoints as estimated by CIBERSORT, stratified for TRs and NRs. Each dot is a sample coloured for TRs (red) or NRs (blue).

We observed another layer of heterogeneity among donors based on pre-vaccination antibody titres. Previous studies have discussed the impact of high pre-vaccination hemagglutination inhibition (HAI) titres that lead to reduced antibody fold change upon vaccination^8, 20–22^. Within our cohort, approximately one-third of the donors (H1N1:36%, B:34%, H3N2:33%) followed this pattern. Nevertheless, the majority of responders in this cohort (TRs and Others) still showed a high antibody response to vaccination that was independent of pre-vaccination titres, suggesting that pre-vaccination titres are not the only variable defining the vaccine response (Figure 1B). Consistent positive correlations among antibody titres against the three influenza strains and positive correlations between HAI titres and microneutralization (MN) titres per strain were observed (Supplementary Figure 1A-1B). We did not observe statistically significant differences in serological responses between elderly males and females (Supplemental Figure 1C). Across two seasons, we observed a significant negative correlation of antibody titres with age for the H1N1 strain, but not for the H3N2 and B strains (Supplementary Figure 1D). These observed patterns were replicated in the smaller cohort.

### Distinct transcriptional signatures between elderly triple and non-responders

We first evaluated transcriptome data generated from the 10 highest (TRs) and 10 lowest (NRs) responders to identify transcriptional differences between the groups and the transcriptomic dynamics of vaccine response resulting in 1466 significantly differentially expressed genes (DEGs) (adj. p-value < 0.01, Figure 1C). Functional significance of the differential transcriptome profiles between TRs and NRs (independent of time) was identified using gene set enrichment analysis (GSEA) with blood transcriptome modules (BTMs)^23^ as background (Supplementary Data 1). We found that the top 40 most significantly enriched modules showed a consistent upregulation of T and B cell related, and T cell activation modules in TRs, whereas NRs showed upregulation of inflammatory and inflammatory signalling modules (Figure 1D). These pathways were consistently upregulated in TRs and NRs at all time points, including pre-vaccination. These differences were also reflected by different proportions of immune cell populations in these donors using transcriptome deconvolution (CIBERSORT^24^, Figure 1E and Supplementary Figure 2A). In line with our enrichment results, we found that TRs had higher proportions of T and B cells, whereas NRs showed elevated proportions of neutrophils. These results agree with reported flow cytometry results^7^ and with the observed trend of higher neutrophils in NRs at the pre-vaccination time point (Supplementary Figure 2B). Despite enrichment of monocyte-related BTMs in NRs, we did not observe differences in monocyte proportions between TRs and NRs in deconvolution or cell blood counts. This suggests higher activity of monocytes in NRs irrespective of cell proportions (Figure 1E and Supplementary Figure 2B).

We next evaluated the pre-vaccination BTMs alone that separate TRs from NRs. We found TRs showed enrichment for T cell activation and signalling, Th2 differentiation and plasma/B cell modules (Figure 2A, Supplementary Data 2). A recent study described a high inflammatory pre-vaccination endotype that is predictive of high vaccine responders in adults, but not in the elderly^13^. In line with these results, we did not observe enrichment of inflammatory BTMs at the pre-vaccination time point in TRs. Instead, we observed an opposite pattern, in which TRs had lower enrichment of inflammatory modules compared to NRs (Supplementary Figure 3A). When we evaluated the expression patterns of the genes enriched in these pathways, we found only CD83 and IL23A to be significantly upregulated in TRs at all time points, suggesting that only select genes are important in elderly TRs and support the immune response (Supplementary Figure 3B). While we observed a gene expression pattern associated with cells engaged in humoral responses in TRs (which is in line with their high responsiveness to vaccination), the NRs presented a consistently upregulated inflammatory signature.

**Figure 2.**
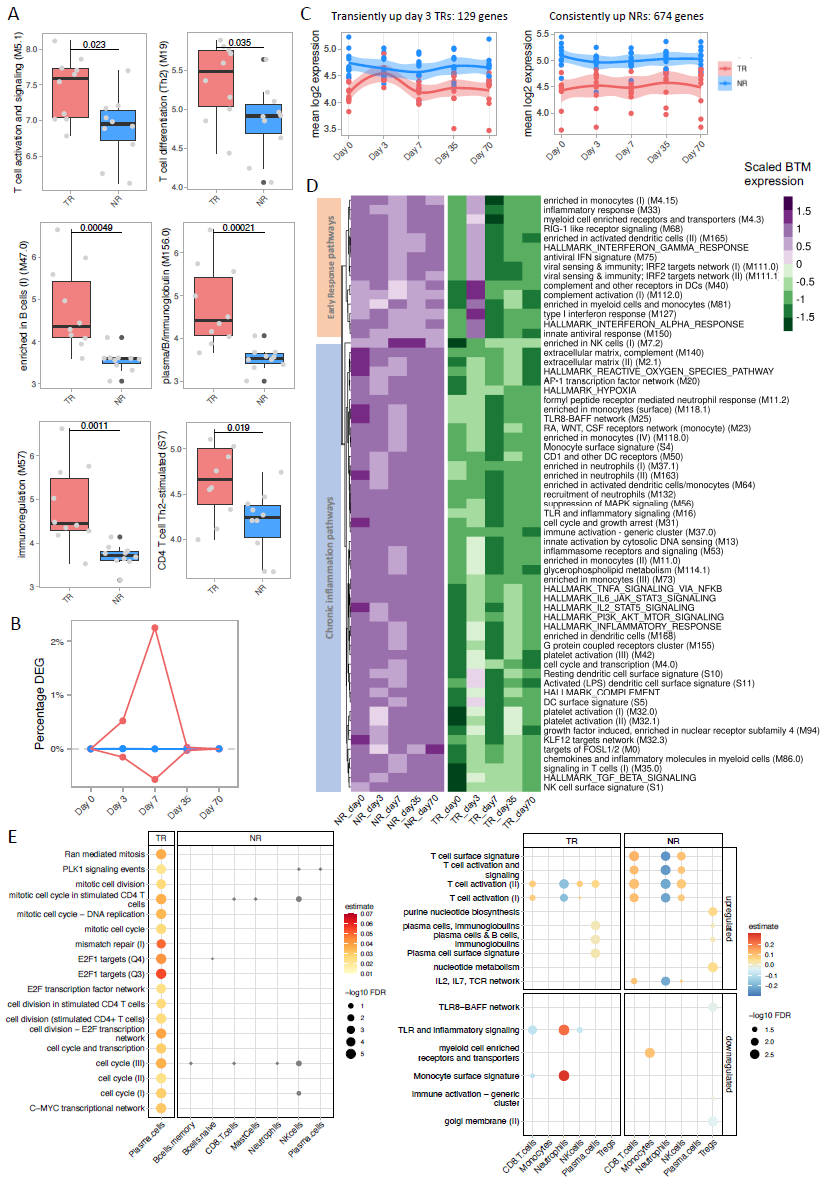
Low inflammatory status at pre-vaccination stage indicator of high responders. **(A)** BTMs significantly upregulated in TRs at the pre-vaccination timepoint. Boxplots show median with interquartile range and Tukey whiskers. **(B)** Line plot summarising changing gene expression in TRs (red) and NRs (blue) upon vaccination indicated by the percentage of significantly differentially expressed genes over time compared to pre-vaccination. **(C)** Line plots for two clusters of genes, where the cluster "Transient day 3 TRs" contains genes upregulated in TRs at day 3 post-vaccination. Genes in the cluster "consistent up NRs" are upregulated in NRs at all timepoints. Each dot represents the mean of genes per donor for each cluster, line plot stratified for TR and NRs. **(D)** Mean expression of genes for significant BTMs and HALLMARK pathways clustered shows split based on transient upregulation in TRs (top) and downregulation in TRs at all time points (bottom). **(E)** Association of CIBERSORT cell proportions with cell cycle BTMs (left) and cell type signature BTMs (right) at seven days post-vaccination. Size of the dots represent adjusted p-values from linear mixed models, whereas colour represents the estimate from the model. Associations with adjusted p-values < 0.05 plotted, for TRs, while all associations for NRs with cell cycle BTMs only irrespective of adjusted p-value threshold (grey). Associations with adj. p-value < 0.05 for cell type signature BTMs for NRs shown.

### Major differences in post-vaccine transcriptional dynamics between elderly triple and non-responders

We then assessed transcriptional changes resulting from vaccination over time, in each group separately. Based on significant DEGs at each time point compared to pre-vaccination, we observed distinct post-vaccination dynamics between TRs and NRs. TRs showed an early response at three days post-vaccination, evident from a small proportion of DEGs, followed by the highest change in gene expression at day seven before going back to pre-vaccination gene expression levels. However, NRs did not show significant DEGs at any time point post-vaccination compared to pre-vaccination (Figure 2B and Supplementary Figure 3C). Interestingly, ∼18% of genes significantly upregulated at day three in TRs were also significantly upregulated in NRs at all time points (Figure 1C and 2C). Functionally, the day three transcriptomic profile was enriched in innate immune modules related to antiviral, interferon and complement activation (Supplementary Figure 3D, Supplementary Data 3). Consequently, the upregulated inflammatory genes in NRs were categorised in two clusters depending on their expression patterns: "Transiently up day 3 TRs" or "Consistently up NRs" (Figure 1C and 2C). While “Transiently up day 3 TRs” genes were involved in antiviral response, the "Consistently up NRs" genes were enriched for modules related to AP-1 transcriptional network, neutrophils and other inflammatory signalling modules, which may suggest a chronic inflammatory status in NRs (Figure 1C, 2D). These results highlight that TRs mounted an immune response evident from transcriptional changes at different time points (percentage of DEGs at day three and day seven) post-vaccination. However, NRs maintained an inflammatory profile at all time points, which in turn may impede mounting the vaccine response.

### Elderly non-responders show higher activation of NK cells seven days post vaccination

We next examined day seven transcriptional changes in TRs and NRs separately in greater detail, as day seven showed the highest change in gene expression in TRs compared to pre-vaccination (Figure 2B). GSEA results in TRs showed strong positive enrichment of multiple modules particularly related to cell cycle, plasma cells, immunoglobulins and T cell activation, among others. Even though NRs did not show statistically significant DEGs seven days post vaccination compared to pre-vaccination, GSEA at this time point showed very few enrichments related to cell cycle, T cell activation and plasma cells. However, the inflammatory modules were downregulated in both groups compared to pre-vaccination (Supplementary Figure 3E, Supplementary Data 3).

We associated the enriched modules with cell proportions identified through transcriptome deconvolution. Cell cycle modules were strongly associated with plasma cells in TRs (FDR < 0.05) suggesting clonal expansion of plasma cells, whereas cell cycle modules in NRs did not show significant associations with any cell subsets (Figure 2E, left). Transcriptional changes, particularly in plasma cells and B cells seven days post vaccination have also been shown through single-cell studies^25^. Plasma cell proportions transiently increased at this time point for TRs, whereas this was not seen in NRs (Figure 1E). This observation is supported by previous studies which showed that an increase in antibody secreting cells at day seven correlates positively with high antibody fold change^26^. Instead, activation modules were strongly positively associated with NK and CD8 T cells in NRs. We also observed similar associations in TRs, but they were weaker than in NRs (Figure 2E). The deconvolution results also show higher proportions of activated NK cells in NRs (Figure 1E). Flow cytometry on donors from the replication cohort also confirmed these findings (Supplementary Figure 4). A higher proportion of NK cells in NRs compared to TRs has also been reported in the single-cell dataset^7^. Increased NK cell proportions and activity in the elderly compared to younger adults has been previously reported^8^. In addition, a significant negative association of neutrophils with T cell activation BTMs in NRs suggests modulation of T cell activation by neutrophils. Finally, in NRs, Tregs were negatively associated with immune activation BTM, which may suggest attempts at suppression of inflammation in response to vaccination. Altogether, the observed associations link cell subsets to the functional changes characterised through BTMs and highlight multiple differences in TRs and NRs post-vaccination. Particularly, we observe higher NK cell proportions and higher activation of NK cells as a characteristic of the elderly NRs.

### Post-vaccination plasma proteome and metabolome differences between triple and non-responders highlight key molecules involved in vaccination response

We next evaluated the proteome (Olink-Explore 384 panel) profile between all TRs (n=71) and NRs (n=10) in our cohort, measuring up to 311 proteins. Differential abundance testing between TRs and NRs irrespective of time showed higher CLEC7A in TRs in the discovery cohort (adj. p = 0.03, moderated t-test,), but not in the replication cohort possibly because of the small effect size (Supplementary Figure 5A, 5B). We next examined protein dynamics induced by vaccination separately in TRs and NRs by comparing each time point to pre-vaccination. Unlike the distinct dynamics observed at the transcriptome level in TRs and NRs, changes in protein abundance in both TRs and NRs seven days post-vaccination were observed (Figure 3A, Supplementary Data 4). In TRs, 14 proteins were upregulated at seven days post-vaccination (Figure 3B), among which TNFRSF13B (also known as TACI) has been widely described for its role in B cell proliferation and plasma cell differentiation^20, 27, 28^. Other proteins upregulated in TRs include IFNLR1^29, 30^, a receptor for type three interferons; CCL3^31^ or MIP1a, CD48 and SLAMF7. TNFRSF13B, IFNLR1 and CCL3 also showed a trend of increased abundance in the replication cohort at day seven post-vaccination (Supplementary Figure 5B). On the other hand, five distinct proteins were upregulated in NRs at day seven post-vaccination, including PARP1, EGLN1 and SERPINB8. PARP1 plays a role in oxidative stress-induced inflammation^32^ and also in recruiting NK cells in a viral response^33^. EGLN1/PHD2 and SERPINB8 are involved in anti-inflammatory roles^34, 35^. At day 35, we found relatively fewer changes (Supplementary Figure 5C). Altogether, differences in plasma proteome dynamics in TRs and NRs highlights the differential response to vaccination induced in the two groups.

**Figure 3.**
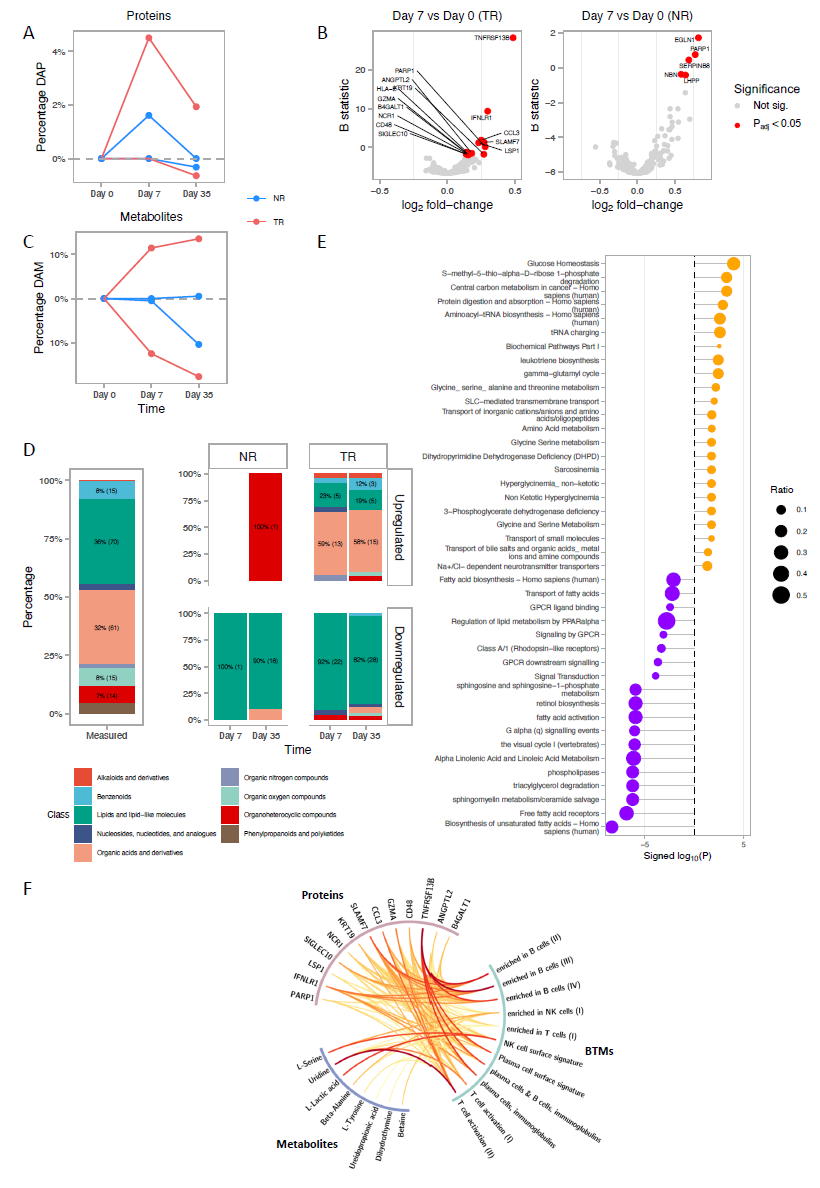
Plasma proteins and metabolite changes upon vaccination are distinct in TRs and NRs. **(A)** Line plot showing significant changes (padj < 0.05) in protein abundance in TRs (N = 71) and NRs (N=10) post-vaccination. TRs show a higher percentage of significantly regulated proteins, while NRs show a comparably dampened response. **(B)** Volcano plot of proteins upregulated in TRs and NRs 7 days post vaccination with significant proteins coloured in red and labelled; Proteins were considered significant at adj. p-value < 0.05 **(C)** Line plot showing significant changes in metabolite abundance in TRs (N=71) and NRs (N=10) post vaccination. (D) Significantly differentially regulated metabolites categorised based on their taxonomic class as annotated by the Human Metabolome Database (HMDB). The upper and lower parts of the bars represent up- and down regulated metabolites, respectively. The "Measured" column represents the total universe of metabolites that we considered (192 endogenous metabolites, see Methods). The TR and NR columns indicate significantly regulated metabolites at each timepoint compared to day 0. **(E)** Pathways enriched for metabolites with increased or decreased abundance 7 days post vaccination in TRs. Enrichment of pathways was calculated using IMPALA. Pathways with adj. p-value < 0.05 are shown. **(F)** Integration of proteins and metabolites with increased abundance at 7 days post vaccination with transcriptome pathways in TRs. Linear mixed models were used to estimate the association between protein / metabolite abundance and mean transcriptomic pathway activity (Methods). Drawn links are significant with adj. p-value < 0.05. Colour and width for the links are based on p-values, where wide red links indicate increased significance.

We also assessed untargeted metabolomic data measured by liquid chromatography-gas spectrometry (LC-MS) and focused specifically on 192 endogenous metabolites (see Methods). No significant differentially abundant metabolites between TRs and NRs were found, irrespective of the time points. However, metabolites in TRs showed significant changes at day seven and 35 compared to pre-vaccination, whereas this was less evident in NRs (Figure 3C top, Supplementary Data 5). Summarising the metabolites into taxonomic classes, TRs showed significant increased abundance of organic acids, particularly amino acids, crucial for adequate immune function both at seven and 35 days post-vaccination^14^. On the other hand, metabolites with reduced abundance both at seven and 35 days post-vaccination were lipids and lipid-like molecules, more specifically fatty acyls (Figure 3C bottom, Supplementary Figure 5D, 5F). These findings were validated in the replication cohort, suggesting that overlapping metabolic signatures were induced by vaccination in both seasons (Supplementary Figure 5E). Interestingly, four out of five upregulated lipids and lipid-like molecules in TRs are bile acids. Bile acid perturbation has been described in antibiotics-treated donors with low antibody response to vaccination^12^. In contrast, NRs did not show any significant differences in metabolite abundance seven days post-vaccination, but instead showed a decrease in lipid abundance 35 days post-vaccination (Figure 3C top and bottom). Pathway over-representation analysis in TRs at day seven showed significant upregulation of metabolic pathways involving amino acid, glycine/serine metabolism and glucose homeostasis (Figure 3E, Supplementary Data 6). On the other hand, downregulated metabolic pathways were related to fatty acids, linoleic acid metabolism and signal transduction, among others. In summary, the increased and sustained abundance of amino acids with reduced abundance of fatty acyls over time suggests the importance of these metabolite classes in vaccine response.

Furthermore, we integrated plasma molecules upregulated at day seven with transcriptome BTMs to derive their functional significance. In TRs, protein concentrations of TNFRSF13B and IFNLR1 were strongly positively associated with B cells and plasma cell immunoglobulin modules from transcriptome (False Discovery Rate (FDR) <0.05). The CCL3, GZMA and CD48 protein concentrations were also significantly positively associated with T cell transcriptional modules, suggesting their role in the activation of T cells. Among metabolites, uridine and tyrosine also showed positive associations with T cell activation and plasma cell numbers, which may imply their involvement in mounting the immune response. Uridine has previously been shown to be important for T cell proliferation^36^. Beta-alanine, lactic acid and serine all positively correlated with NK cells (Figure 3F). In contrast, no significant associations between the proteome and transcriptional modules were found in NRs. However, EGLN/PHD2, SERPINB8 and NBN were negatively associated with inflammatory pathways (nominal p-value < 0.05, Supplementary Figure 6). Combining these results, we show distinct changes in the proteome and metabolome following vaccination and describe their potential significance in shaping the vaccination response in the elderly vaccine responders.

### Multi-omics integration reveals a common axis of variation across layers separating high and low responders

Next, we integrated the three omics layers of the highest (TRs, n=10) and lowest responders (NRs, n=10) to examine the interconnected processes underlying the heterogeneous influenza vaccine response in the elderly. Using multi-omics factor analysis (MOFA)^37^, an unsupervised method, 15 latent factors were identified which explained different levels of data variation along with varying contributions of omics layers within each latent factor (Figure 4A). Among these, factor three showed a common axis of variation among three omics layers that separated TRs from NRs irrespective of time (Figure 4B). Examining significance of the other factors showed that factors four and six were consistent with the time dynamics differences observed in TRs and NRs (Supplementary Figure 7A).

**Figure 4.**
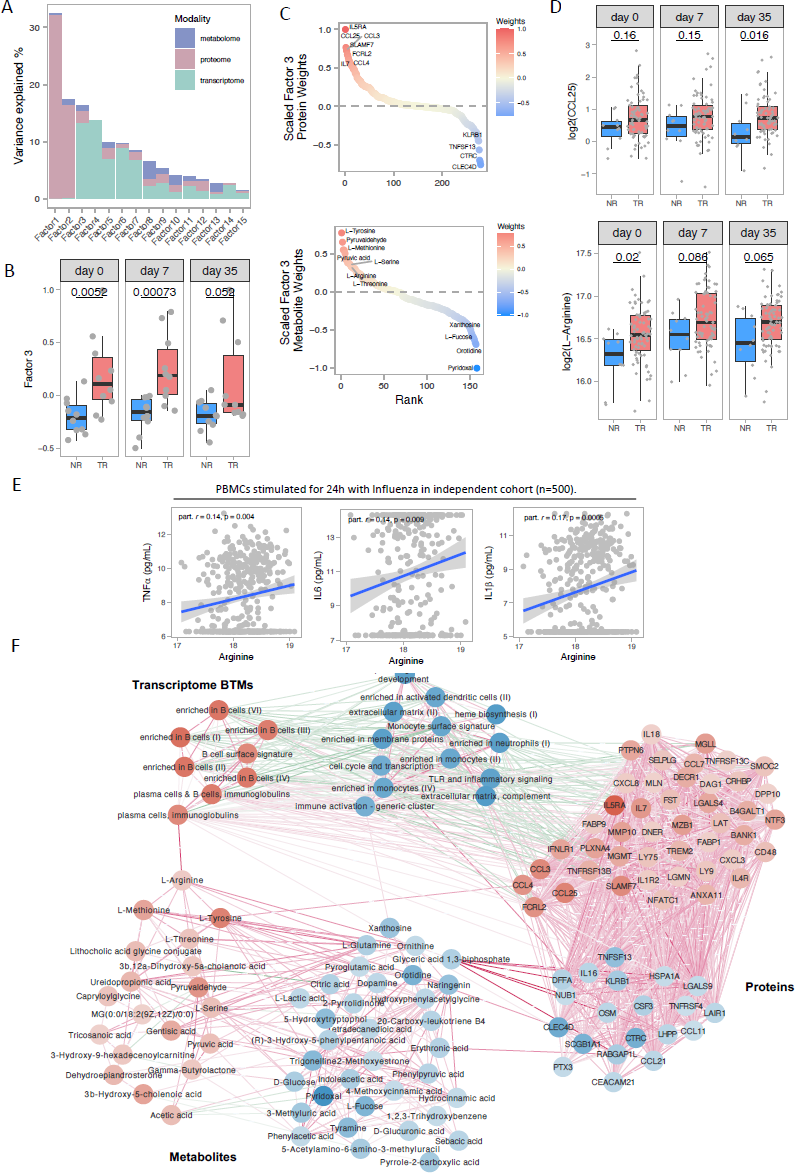
Multiomics integration of high and low responders. **(A)** Bar plot showing the proportion of explained variance per factor resulting from unsupervised factor analysis method MOFA. Colours within each bar indicate the contribution of each modality. **(B)** MOFA Factor 3 separates TRs from NRs irrespective of time and across modalities. Y-axis represents the Factor 3 variance value attributed to each donor. P-values are generated using a Wilcoxon ranked-sum test. **(C)** Top most positive and negative molecules within the metabolome and proteome modalities as calculated by MOFA. Molecules are ranked on MOFA scaled weights for factor 3. Positive weights are for TRs, whereas negative weights correspond to NRs. **(D)** Abundance of CCL25 and L-arginine in all 81 TR and NRs. P-values are generated using a Wilcoxon ranked-sum test. **(E)** Association of arginine with cytokine response post influenza stimulation in 500 FG cohorts. P-values are generated using t-test. Partial correlation estimate was calculated, corrected for age and gender **(F)** Integrative network of transcriptome, proteome and metabolome data. Nodes in the network correspond to molecules (proteome, metabolome) or BTMs (transcriptome). Only molecules and pathways identified by MOFA weights (Supplementary Figure 6F) are plotted. Edges in the network (all adj. p-value < 0.05) are statistical associations from linear mixed models (see Methods), where red edges represent positive associations and blue edges represent negative associations.

To examine the significance of this common axis (latent factor three), we examined the top proteins and metabolites identified through this factor (Figure 4C) and evaluated these in all TRs (n=71) and NRs (n=10). Among the proteins, CCL25 and CCL3 were consistently more abundant in TRs than in NRs. Similarly, for metabolites, the amino acids arginine and methionine were more abundant in TRs (Figure 4D and Supplementary Figure 7B). To further assess the significance of these amino acids in immune function, we examined the association of amino acids with cytokine production capacity, particularly induced upon influenza stimulation in an independent cohort of 500 healthy individuals^38, 39^. Arginine and methionine showed significant positive associations with IL1β, IL6 and TNFL (Figure 4E, Supplementary Figure 7C). The increased abundance of these amino acids in TRs and their positive association to pro-inflammatory cytokine production may indicate their involvement in shaping the adaptive immune response to influenza vaccination via modulation of cytokine production.

Finally, we then created a network of significant intra- and inter-omic connections by associating plasma molecules with transcriptome BTMs (Methods), overlaying the weights assigned for TRs (red) and NRs (blue) from MOFA latent factor three (Figure 4E). Across omic layers, the transcriptome and proteome were highly connected, whereas the metabolome was less connected to the other two layers. The metabolite arginine was positively associated with TR-related BTMs involving plasma cells and immunoglobulins. Similarly, xanthosine was positively associated with immune activation BTMs, which was estimated as a predictor for NRs. Both xanthosine and orotidine are involved in nucleotide metabolism which plays an important role in inflammation^40, 41^. We also observed strong positive associations of glyceric acid 1,3-bisphosphate/1,3-BPG with up to seven proteins that are predictors for NRs. In summary, we describe a common axis of variation in transcriptome, proteome and metabolome which highlights the interconnected differences in influenza vaccine response between elderly TRs and NRs.

### Pre-vaccination high plasma IL-15 concentration negatively correlates with antibody response

One of the prerequisites for complete protection is the ability to produce sufficient antibodies against all three strains of the influenza vaccine. To prioritise the important pre-vaccination plasma proteins for antibody response to three strains of influenza vaccine concurrently, we used partial least square regression (PLSR) in 200 donors (discovery cohort). This allowed us to perform a systematic investigation of heterogeneity at the pre-vaccination time point. The resulting proteins from the top three PLSR components explain ∼40% variance of the antibody fold change within the cohort (Figure 5A). We evaluated the top 30 proteins from each of the three components to examine the directionality of the association with antibody response per strain (Figure 5B). Overall, the majority of the prioritised proteins showed contrasting associations with the three strains. Interestingly, TNFSF13/APRIL and IL-15 were negatively associated with antibody fold changes for all three strains (Figure 5B,5C and Supplementary Figure 8A left). Both these proteins have been described as pro-inflammatory cytokines and in the pathogenesis of influenza infection^42–44^. In the replication cohort, we also found IL-15 negatively associated with high antibody changes for two strains, whereas this was not true for TNFSF13, perhaps due to the small effect size that is hard to replicate in a smaller cohort (Supplementary Figure 8A right, 8B). Single-cell RNA sequencing on human PBMCs showed that IL-15 is mainly expressed by monocytes^45^ (Supplementary Figure 8C). Based on these results, the contrasting relationship of the majority of prioritised proteins against the three strains suggests a complex relationship of these proteins involved in providing complete protection against influenza infection. However, the consistent negative association of IL-15 with antibody response in both the discovery and replication cohort suggests a detrimental role in protection against influenza.

**Figure 5.**
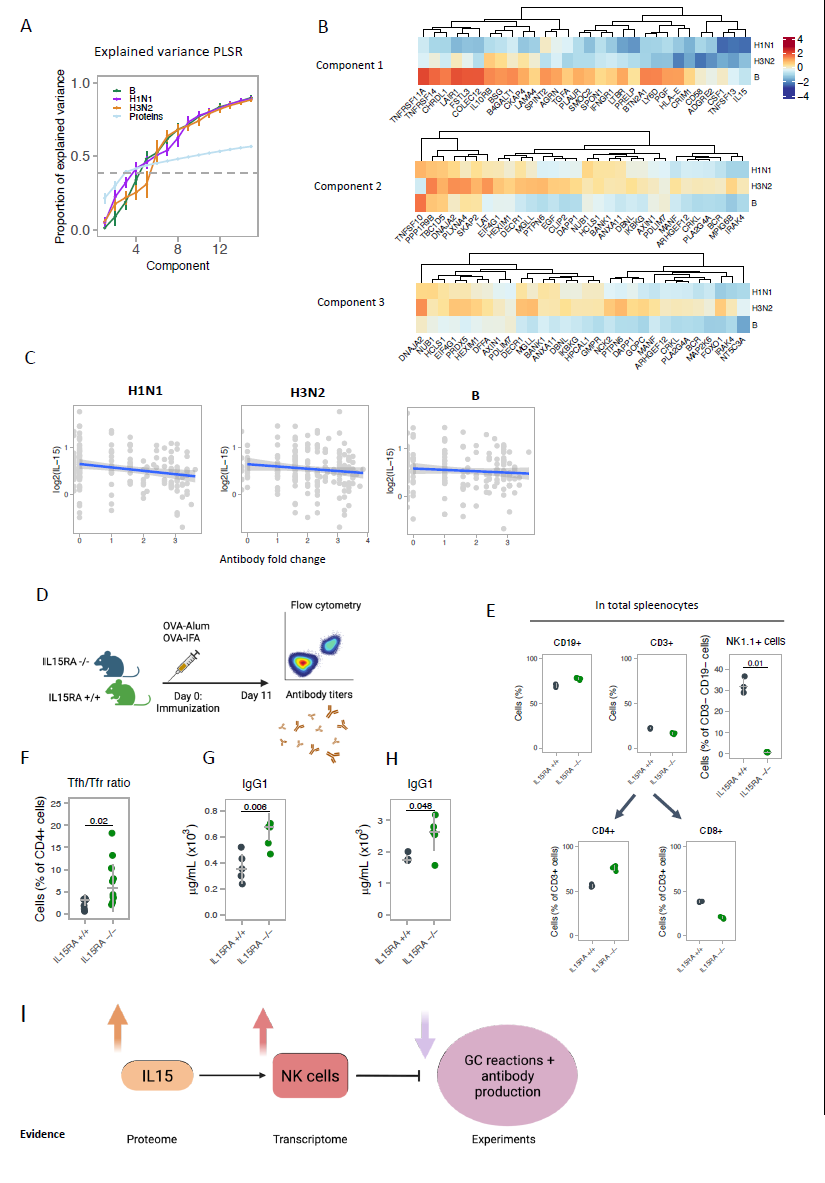
Pre-vaccination plasma proteome correlates to the antibody response to vaccination. (**A**) Proportion of co-variation in proteome and antibody fold change explained by each component for either each strain or the predictors examined through PLSR analysis with 10-fold cross validations (N = 200). The first three components using the predictors were able to explain ∼ 40% co-variation in the antibody response and pre-vaccination protein abundance (**B**) Heatmap of t-test statistics for top predictors with antibody fold change for top three components as calculated using rank product analysis for these components. Most proteins show heterogenous associations with the antibody fold change against the three influenza strains, with notable exceptions for IL-15 and TNFSF13 in component 1. (**C**) Correlation plots of IL-15 against antibody fold change for all 200 donors. (**D**) *IL15RA*^+/+^ and *IL15RA*^−/−^ mice were immunised with OVA-Alum or OVA-IFA in the footpad, followed by analysis of antibody titres, and lymphocyte populations from inguinal lymph nodes 11 days post-immunization. (**E**) Proportion of B (CD19+), T (CD3+), NK (NK1.1+) cells, as well as CD4+ and CD8+ T cell subsets, among splenocytes of unimmunized *IL15RA*^+/+^ and *IL15RA*^−/−^ mice. (**F**) Ratio of Tfh/Tfr cells, calculated from their frequency among total CD4+ cells, within LNs of immunized *IL15RA*^+/+^ and *IL15RA*^−/−^ mice. (**G**) Serum concentration of OVA-specific IgG1 in mice immunized with OVA-alum, or (**H**) immunized with OVA-IFA. (**I**) Schematic summarising the mechanism of IL-15-mediated activation of NK cells leading to suppression of germinal centre responses and low production of antibodies.

### High IL-15 concentration leads to reduced antibody titres through NK cells mediated suppression of germinal centre responses

IL-15 is important for NK cell proliferation and maturation^46–48^. From both bulk and previously reported single-cell transcriptomics^7^, we observed higher frequencies of activated NK cells and total NK cell numbers in NRs (Figure 1E). NK cells have been described to suppress germinal centre (GC) responses^49, 50^. We therefore examined the impact of a perturbation of the IL-15-NK cell axis on antibody production with *IL15RA*-deficient mice. We immunised *IL15RA*^+/+^ and *IL15RA*^−/−^ mice with ovalbumin (OVA-IFA or OVA-Alum) to examine the resulting humoral response 11 days post-immunization, at the time of maximum GC-response in this animal model^51^ (Figure 5D). *IL15RA*^−/−^ mice showed a marked reduction of NK cells in the spleen (as anticipated, given the importance of IL-15 for NK cell numbers), without significant differences on B cell populations, but with slight differences on the frequency of CD4+ and CD8+ T cells (a population that can also be affected by IL-15 reduction) (Figure 5E, Supplementary Figure 9A). In the draining lymph node, we observed a consistent increase of Tfh cells and a decrease of Treg and Tfr cells in *IL15RA*^−/−^ mice, although without reaching statistical significance (p-values > 0.05) (Supplementary Figure 9B). However, the Tfh/Tfr ratio, critical in the regulation of GC responses^52^, was significantly higher in *IL15RA*^−/−^ mice (p-value = 0.02, Wilcoxon ranked sum-test, Figure 5F). In the same mice that had an increased Tfh/Tfr ratio, we observed higher production of antibodies, across two different immunisation protocols (p-values = 0.006 (OVA-Alum) and 0.048 (OVA-IFA), Wilcoxon ranked-sum test, Figure 5G-H). In conclusion, these results suggest that the reduction of IL-15 and NK cells led to more effective antibody production, in line with the data from elderly NRs who displayed higher IL-15 and NK cell activation together with poor antibody response (Figure 5I).

### Pre-vaccination plasma malic and citric acid concentrations negatively correlate with antibody response

To evaluate the pre-vaccination metabolite levels with antibody responses against three strains, PLSR was used in a similar way as described above. Herein, the metabolites in the top eight PLSR components explain ∼40% variation of the antibody fold change among donors suggesting high inter-individual variation in the donors as a consequence of metabolic differences (Figure 6A). Metabolites prioritised in the first component showed malic acid and citric acid as top molecules, with negative associations to antibody response (Figure 6B, 6C and Supplementary Figure 9A). To derive the role of citric acid and malic acid in immune function, we associated pre-vaccination abundance of these metabolites with deeply phenotyped immune cell counts and cytokine production induced upon influenza stimulation in two independent cohorts of 300 and 500 healthy individuals, respectively^38, 39, 53, 54^. Citric acid showed significant negative associations with CD4+CCR6+CCR5+CCR7+ T cells along with three different subsets of immature neutrophils (FDR<0.05) (Supplementary Figure 9B). Malic acid showed strong negative associations with TNFL, IL6 and IL1β cytokine levels upon influenza stimulation (Figure 6D). Furthermore, we found betaine and cysteine as the top predictors from components one and two, respectively, to be consistently positively associated with antibody response across all strains (Figure 6B and Supplementary Figure 9C). Betaine has been described for its inhibition of IL1β production and release^55, 56^ and for its suppression of pro-inflammatory signalling during ageing^57^. These results reflect the impact of pre-vaccination metabolite abundance linked to different physiological immune states across responders culminating in differential vaccination response in the elderly.

**Figure 6.**
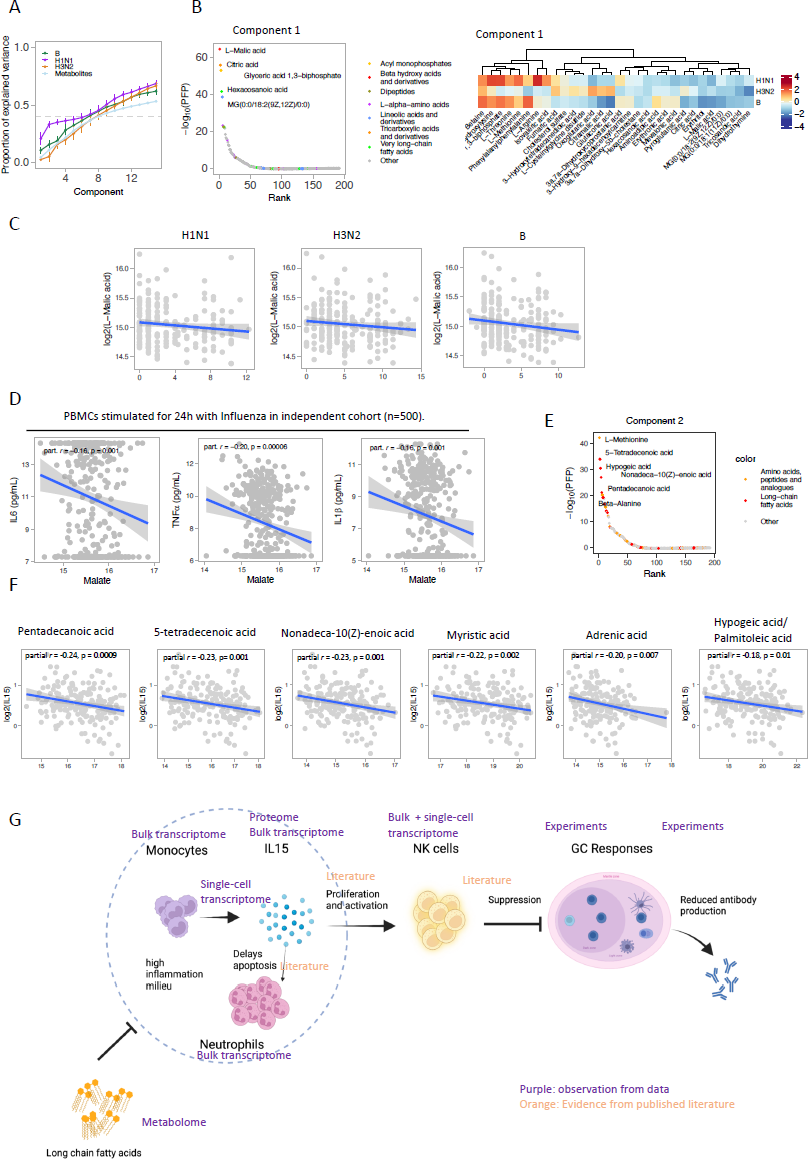
Pre-vaccination plasma metabolites as modulators for vaccination response. **(A)** Proportion of co-variation in endogenous metabolites and antibody fold change explained by each component for either each strain or the predictors examined through PLSR analysis with 10-fold cross validations (N = 200). The first 8 components were able to explain ∼ 40% co-variation in the antibody response and pre-vaccination metabolite abundance **(B)** Plot of metabolites from component 1 with their ranks and estimated percentage of false predictions (PFP) (left), Heatmap of t-test statistic for the top predictors with antibody fold change for the top component as calculated using rank product analysis for these components (right). Most metabolites show heterogeneous associations to the different influenza strains in the vaccine, except for malic acid and citric acid showing negative association while betaine shows positive association. **(C)** Correlation plots of malic acid against antibody fold change for all three influenza strains for all 200 donors. **(D)** Association of malic acid to cytokine production upon influenza stimulation in an independent cohort of 500 younger healthy individuals, showing a negative trend. P-values are generated using t-test. Partial correlation estimate was calculated, corrected for age and gender **(E)** Top metabolites for component 2 with their ranks and estimated percentage of false predictions shows long chain fatty acids as top candidates. **(F)** Negative association of these long chain fatty acids identified in PLSR component 2 with IL-15. P-values are generated using t-test. Partial correlation estimate was calculated, corrected for age and gender **(G)** Schematic describing the role of IL-15 in delayed neutrophil apoptosis, maturation and activation of NK cell populations, leading to suppression of GC responses and reduced antibody production and a chronic inflammatory status. Evidence for each observation is derived through different omics layers and experiments. Connections among different observations were also supported by published findings.

### Unsaturated long chain fatty acids are negatively associated with pre-vaccination IL-15 abundance

Given the putative role of high IL-15 in suppressing antibody production in the elderly, we evaluated the relationship of pre-vaccination IL-15 abundance and metabolites. Pre-vaccination IL-15 showed strong negative associations with odd-chain (pentadecanoic acid) and certain unsaturated long-chain fatty acids (LCFAs, e.g. palmitoleic acid) (p value < 0.02) (Figure 6F and Supplementary Figure 9D). These LCFAs were also among the top 30 predictors for PLSR component two, explaining variation in antibody fold change among the donors (Figure 6E and 6F). LCFAs and especially poly-unsaturated fatty acids have been described as important immunomodulatory metabolites to protect against infections^58^. To test the significance particularly of LCFAs in suppressing inflammatory proteins, the relationship of all pre-vaccination lipid and lipid-like molecules (taxonomy class) with these proteins was examined. LCFAs were negatively associated with inflammatory proteins, whereas other lipid molecules showed both positive and negative associations (Supplementary Figure 9E). We also examined the role of LCFA in a younger cohort (500FG^38, 39^, with 98% donors less than 65 years of age), however, the LCFA-specific negative associations with inflammatory proteins were absent (Supplementary Figure 9E). Among the LCFAs, the most significantly negatively associated metabolite was pentadecanoic acid, an odd-chain fatty acid with anti-inflammatory properties and resistance to oxidation^59, 60^. Taken together, the data suggests an age-dependent role of LCFAs in the control of inflammation. We described different metabolites associated with IL-15 and other inflammatory proteins, as well as the antibody fold change, suggesting their role in modulating the vaccine response in the elderly, particularly LCFAs as potential pre-vaccination suppressors of high inflammation.

## DISCUSSION

In this work, we evaluated a large cohort of healthy elderly individuals to understand the heterogeneity of influenza vaccination response, specifically to address two main questions:

1. How does the vaccination response differ between responders and non-responders, and
2. Which pre-vaccination biomarkers correlate with the vaccination response. To answer these questions, we examined the transcriptome, proteome and untargeted metabolome of these donors separately, as well as across omics layers.

In the transcriptome, proteome, and metabolome layers, we observed post-vaccination dynamics that were indicative of mounting of an immune response to vaccination in TRs whereas, as expected, these dynamics were absent or altered in NRs. NRs were instead characterised by increased and activated NK cell populations and persistent inflammation. In line with these findings, the NK cell receptor *KLRB1* was shown as a negative predictor for vaccine response in the elderly^21^, whereas *NKG2C* expression was reported as positively correlated with influenza vaccination^61^ in healthy individuals. This opposing observation of NK cells may be related to the ageing immune system. Signalling through inflammatory molecules is important for mounting an immune response, while persistent low-grade inflammation, termed ‘inflammaging’, can result in exhaustion and is detrimental to the vaccine response^62, 63^. In NRs, evidence from the transcriptome suggests persistent inflammation driven through AP-1 transcriptional networks, among others. On the other hand, the post-vaccination proteins consist of proteins with anti-inflammatory roles. These opposing findings from the different layers may suggest a dysregulation where NRs are attempting to suppress the inflammation resulting from vaccination required for mounting the necessary immune response. A recent study has revealed endotypes predictive of antibody response to vaccination across different vaccines, including influenza^13^. However, this endotype was not identified in elderly (>50 years) individuals. Our study extends these findings in the elderly and defines the pre-vaccination transcriptome profile separating high and low responders, with adaptive immune modules enriched in high responders. This suggests that a high persistent inflammatory status predictive of high responders in the young is increasingly detrimental with age.

Using multi-omics integration for day seven and across time points, we were able to find key molecules that are important in supporting and leading to a productive vaccine response. Interestingly, some of these identified molecules e.g. CCL25, CCL3 and arginine have already been studied as potential immune-adjuvants in vaccination and/or as supplementation for a better humoral response to vaccination^31, 63–65^. CCL25, a chemokine ligand for CCR9 has been shown to play both pro and anti-inflammatory roles in various diseases^66^. CCL3 and its receptor CCR5 have been implicated in leukocyte recruitment into the airway leading to upregulation of antiviral responses in influenza infection^67^. Amino acids in general are crucial for immune function^14, 15^. In fact, we observed consistent upregulation of amino acids post-vaccination in TRs, but not in NRs. In this study, we showed functional associations of plasma concentrations of metabolites arginine and methionine to cytokine production upon influenza stimulation using an independent cohort. The upregulation of these amino acids in TRs irrespective of vaccination and the role of these molecules in cytokine production suggests an important role in protective immunity and consequently markers of protection in elderly TRs. In summary, we indicate biomarkers that support both a robust immune response to influenza stimulation and the vaccine response.

Pre-vaccination plasma IL-15, malic acid, and citric acid concentrations are negatively associated with antibody response. We propose a model where high abundance of pre-vaccination IL-15 leads to increased activated NK cells, which in turn inhibit the GCs to suppress antibody production (Figure 6F). Modulation of adaptive immune responses through NK cells has been proposed for better vaccine design, where NK cells control the magnitude and quality of immune response^68–70^. Previous work and our current results have shown lower levels of T and B cells in NRs compared to TRs^7^. The simultaneous decrease of T and B cells, with increased NK cell proportions may further cause the suppression of antibody responses in NRs. Monocytes are the main source of IL-15 in immune cell populations in human blood and are reported to produce more IL-15 with increasing age^71^. We did not observe higher monocyte proportions but higher monocyte (inflammatory) transcriptional activity in NRs. IL-15 has also been described to inhibit apoptosis of neutrophils^72^, which were more abundant in our NRs.

Together, these findings suggest contributions from inflammatory monocytes and neutrophils in maintaining a persistent inflammatory milieu in NRs. The receptor for IL-15 is also expressed by dendritic cells and the importance of the cross-talk between NK cells and dendritic cells in immune cell activation and maturation has been established^73^. We did not observe differences in deconvoluted dendritic cell proportions between TRs and NRs. However, cross-talk between these cell subsets within the secondary lymphoid tissues could play a role during influenza infection and its role in vaccination in the elderly is of interest for future research. The top predictors from the metabolome PLSR component two consisted of methionine and fatty acids, particularly unsaturated LCFAs. Methionine was one of the top predictors for TRs picked by MOFA, as well as one of the top metabolites upregulated at day seven post vaccination in TRs. Interestingly, unsaturated LCFAs were all negatively associated with IL-15. Recent work has highlighted the role of fatty acid metabolism in the IgG1 response to influenza^12^. In line with this, we observed consistent downregulation of fatty acyls in TRs post-vaccination, whereas this only occurred in NRs at the later time point (day 35). These findings suggest a multifaceted role of fatty acyls by serving as energy molecules and/or signalling molecules, thereby modulating the immune response. Metabolites with known anti-inflammatory properties such as (poly)-unsaturated LCFAs and odd-chain fatty acids could alleviate the chronic inflammation, leading to a more productive vaccination response in the elderly.

We also need to acknowledge the limitations of this study. Larger cohorts are required in future studies to fully explore the molecular mechanisms leading to a protective or detrimental vaccine response. This includes examining the role of genetics in interindividual variation, the host microbiome and its interactions with the metabolome. The role of comorbidities frequently observed in the elderly (type 2 diabetes, cardiovascular diseases, etc.) may also affect the response to vaccination. Indeed, we found that a history of diabetes or herpes zoster (shingles) were independent risk factors for a poor humoral immune response to the H1N1 vaccine antigen in this study cohort^19^. Furthermore, it was recently identified in a study of responsiveness to COVID-19 vaccination that comorbidities, such as diabetes, are associated with a pro-inflammatory response and reduced vaccine responsiveness^74, 75^. Although “inflammaging” associated with pro-inflammatory cytokines is also emerging as a common feature of poor vaccine responsiveness in people with chronic diseases and old age, it will be critical to understand the potential universal character of the identified mechanisms and biomarkers by performing studies using other vaccines and populations. Therefore, future studies incorporating these factors will provide an improved understanding of the vaccine response in the elderly. Furthermore, *in vivo* experiments and clinical trials are required to assess whether modulation of metabolites can improve the vaccine response in (elderly) non-responders.

## Data Availability

Raw data and code will be made available upon publication. Datasets are fully available for reviewers.

## ACKNOWLEDGEMENTS & FUNDING

The authors thank all the participants in this study. We thank the Research Core Unit Metabolomics of the Hannover Medical School for supporting the targeted metabolomics measurements. This work was supported by an ERC starting Grant (948207), a Radboud University Medical Centre Hypatia Grant (2018) and the Deutsche Forschungsgemeinschaft (DFG; German Research Foundation) under Germany’s Excellence Strategy - EXC 2155 project number 390874280 to YL and iMed, the Helmholtz Association’s Cross Programme Initiative on Personalized Medicine, to CAG and FP. C-JX was supported by a Helmholtz Initiative and Networking Fund (1800167). MGN was supported by an ERC Advanced Grant (833247) and a Spinoza Grant of the Netherlands Organization for Scientific Research. LG research was supported by grants HR22-00741 (“la Caixa” Foundation), and 2022.04903.PTDC (Fundação para a Ciência e Tecnologia Portugal). This project was also partly supported by the European Union’s Horizon 2020 research and innovation programme under the Marie Sklodowska-Curie grant agreement No. 955321

## AUTHOR CONTRIBUTIONS

Conceptualization and study design: YL

Sample collection: ST, PR, FP, CAG

Sample preparation: LZ, AA

Data analysis and investigation: SK, MZ, NN.

Murine experiments: RP, LG

Discussion and interpretation: SK, MZ, ST, PR, JBB, CJX, LABJ, MGN, LG, CAG, FP, YL

Writing - original manuscript: SK, MZ, JBB, CJX, YL

Reviewing and editing manuscript: All authors.

## METHODS

### Study population

The study population has been described before in detail by Riese, Akmatov and colleagues^7, 17^. Briefly, a prospective population-based study across two Influenza seasons (2014/2015 and 2015/2016) was performed among elderly individuals (>65 years of age) from Hannover, Germany. Donors from both seasons did not overlap. Participants were recruited from a random sample from the local population registry and represented the general population as verified by a survey^18^. Intravenous blood samples were drawn before vaccination (day 0) and on day 1, day 3, day 6/7, day 21 & day 70 post-vaccination. Hemagglutination inhibition titres and microneutralization titres were measured as described before^7^. Briefly, we followed the formulation of the Fluad vaccine and used the following antigens for titre measurements in serum: H1N1 A/California/7/09 NYMC-X181 (both seasons), H3N2 A/Texas/50/2012 NYMC-223 (season 2014/2015), H3N2 A/Switzerland/9715293/2013 NIB88 (season 2015/2016), B/Massachusetts/02/2012 NYMC BX-51B (season 2014/2015) and B\Brisbane/9/2014 (season 2015/2016). Sero-conversion was defined as either a post-vaccination titre of ≥ 40 for individuals with pre-vaccination titres < 10, or a 4-fold increase in titre upon vaccination for individuals with pre-vaccination titre ≥10. Individuals were classified as Triple Responders (TR, sero-conversion against all three strains), Non-Responders (NR, no sero-conversion against any strain), or Other (sero-conversion against at least one strain but not all three).

### Bulk transcriptome sequencing

Bulk transcriptome sequencing has been described before^7^. In short, ten TRs and ten NRs were selected based on their serological response to vaccination. We included five time points per individual: baseline, day 1/3, day 6/7, day 21/35 & day 60/70. miRNA and total RNA were purified from whole blood samples frozen in PAXgene™ tubes (BD) using the “PAXgene Blood miRNA Kit” (Qiagen). The detailed procedure was previously described^7^.

### Untargeted metabolomics

We assessed the metabolic profiles of 702 individuals from two influenza seasons. Plasma samples were randomised across plates with respect to vaccine response, sampling time point and sex. Polar metabolites were extracted from each sample with 180 uL 80% methanol in a deep well extraction plate using 20 uL plasma by General Metabolics, Boston, USA. Samples were vortexed for 15 seconds, incubated at 4°C for one hour and centrifuged at 4°C, 3750 RPM for 30 minutes. Metabolome profiles of the sample extracts were acquired using flow-injection mass spectrometry. The method described here is adapted from previously described methods^76^. The instrumentation consisted of an Agilent 6550 iFunnel LC-MS Q-TOF mass spectrometer in tandem with an MPS3 autosampler (Gerstel) and an Agilent 1260 Infinity II quaternary pump. The running buffer was 60% isopropanol in water (v/v) buffered with 1 mM ammonium fluoride. Hexakis (1H, 1H, 3H-tetrafluoropropoxy)-phosphazene) (Agilent) and 3-amino-1-propanesulfonic acid (HOT) (Sigma Aldrich). The isocratic flow rate was set to 0.150 mL/min. The instrument was run in 4GHz High Resolution, negative ionization mode. Mass spectra between 50 and 1,000 m/z were collected in profile mode. 5 uL of each sample were injected twice, consecutively, within 0.96 minutes to serve as technical replicates. The pooled study sample was injected periodically throughout the batch. Samples were acquired randomly within plates.

### Targeted metabolomics

Serum concentrations of amino acids were determined by mass spectrometry, using an HPLC-coupled triple quadrupole mass spectrometer (AP4000, Siex) and the AbsoluteIDQ p180 kit (Biocrates Life Science AG, Innsbruck, Austria) following the manufacturer’s protocols^77^.

### Targeted proteomics

We measured 384 circulating proteins in plasma using Olink’s proximity extension assay (PEA) Explore Inflammation panel^78^ in 702 individuals. In the PEA, oligonucleotide-labelled antibodies (’probes’’) bind the protein of interest. Linking of the probes is triggered by close proximity of two antibodies which limits cross-reactivity. Upon linking, the probe sequence is hybridised and subject to extension by DNA polymerases. The resulting sequence is quantified by real-time polymerase chain reaction. Protein values are expressed as normalised protein expression (NPX) values, a relative value on a log2 scale. Quality control of the raw data was performed by Olink (incubation controls, extension controls and detection controls).

### Immunisation, cell isolation and blood serum collection of IL15RA^−/−^ and IL15RA^+/+^mice

Sex-matched *IL15RA*^−/−^ and *IL15RA*^+/+^ C57BL/6 mice, aged from 10 to 12 weeks, were immunized in the back paw footpad with Ovalbumin (OVA, Ovalbumin EndoFit, Invivogen, #vac-pova) emulsified 1:1 (v:v) with either IFA (IFA, Sigma-Aldrich, #F5506) or Alum (Alu-Gel-S, Serva, #12261-01). Each animal was inoculated with a volume of 50 μl per paw containing 80 μg of OVA (OVA-IFA was injected in two paws while OVA-Alum was injected in one paw per mouse). Mice were sacrificed 11 days after immunisation and the cells from the spleen and inguinal lymph nodes (LNs) isolated. Blood serum from each mouse was also collected at the same time point by cardiac puncture. The mice were bred and maintained under specific pathogen-free conditions at the Instituto de Medicina Molecular (iMM), where the experiments were performed under an animal experimentation authorization granted by Direção-Geral de Alimentação e Veterinária (DGAV), the Portuguese national authority for animal health, and iMM-ORBEA, the Ethic Committee for laboratory animal care at the iMM Rodent Facility in Lisbon.

### Flow cytometry of IL15RA^−/−^ and IL15RA^+/+^mice

Characterization of the lymphoid cell populations in the spleen of *IL15RA*^−/−^ and *IL15RA*^+/+^ C57BL/6 strains was done by flow cytometry with the following antibodies: anti-CD19-FITC (MB19-1, eBioscience; 1:100), anti-CD3-APC (145-2C11, eBioscience; 1:100), anti-CD4-BV510 (RM4-5, BioLegend; 1:400), anti-CD8-eF780 (53-6.7, eBioscience; 1:200), and anti-NK1.1-PECy7 (PK136, eBioscience; 1:100). B and T cells subpopulations present in the inguinal LNs were first pre-incubated with anti-CXCR5-Biotin (2G8, BD Biosciences; 1:50) and subsequently stained with anti-CD19-FITC (MB19-1, eBioscience; 1:100), anti-CD95-PE (Jo2, BD Biosciences; 1:100), anti-ICOS-PerCPCy5.5 (7E.17G9, BioLegend; 1:200), anti-PD-1-PECy7 (J43, eBioscience; 1:100), anti-GL7-eF660 (GL-7, eBioscience; 1:100), anti-CD25-APCeF780 (PC61.5, eBioscience; 1:200), anti-CD4-BV510 (RM4-5, BioLegend; 1:400), and anti-CD44-BV605 (IM7, BioLegend; 1:400) together with Streptavidin-BV711 (BioLegend; 1:100). Intracellular Foxp3 (FJK-16s, eBioscience; 1:100) staining was performed using the Foxp3 Fix/Perm Kit (eBioscience, #00-5521-00) according to the manufacturer’s instructions. The samples were acquired on a BD LSRFortessa cytometer from the iMM Flow Cytometry Facility and further analyzed on FlowJo software (TreeStar).

### Anti-OVA IgG quantification of IL15RA^−/−^ and IL15RA^+/+^mice

Serum anti-OVA mouse IgG1 was quantified by ELISA. 96-well plate wells were coated with 50 μl OVA (Ovalbumin EndoFit, Invivogen, #vac-pova) at 20 μg/ml in PBS overnight (4 °C), washed 3 times with ELISA Buffer (Invitrogen, #88-50620-88) and subsequently blocked with 200 μl of ELISA Buffer for at least 30 minutes. After washing, standards and different dilutions of each serum sample were plated in duplicates in the coated wells for at least 90 minutes. The standard curve was obtained with measurements from known concentrations of Anti-OVA [6C8] IgG1 antibody (AbCam, #ab17293). After washing, 50 μl of anti-mouse-IgG1-HRP (SouthernBiotech, #1070-05) diluted 1:2000 were added to the wells and incubated for at least 45 minutes. Wells were washed, and 50 μL of the substrate solution 3,3′,5,5′-tetramethylbenzidine (TMB Single Solution, Life Technologies) were added per well. The reaction was stopped with 25 μL of sulfuric acid (H_2_SO_4_) according to the colour development of the standards. Lastly, optical density (OD) was read at 450 nm within 30 seconds after stopping the reaction. Quantification of Anti-OVA mouse IgG1 in each sample of blood serum was attained by calculating the concentration of antibody considering the OD values obtained for a dilution that fitted within the OD values of the standard curve (0.2 > OD < 1).

### Flow cytometry of human PBMCs

Cryopreserved PBMCs were thawed, rested for 2 h and incubated overnight with purified anti-human CD28 and CD49d at a concentration of 10 µg/ml. For surface staining, cells were incubated with the antibody mixture prepared in PBS for 30 min at 4°C in the dark. Cells were stained for flow cytometric analysis using the following antibodies: CD3 (BUV395, SK7, Cat.-No. 564001, dilution 1:200, BD (Franklin Lakes, New Jersey, USA)) and CD56 (BV785, 5.1H11, Cat.-No. 362550, dilution 1:100, BioLegend (San Diego, USA)). The samples were acquired at a BD Fortessa flow cytometer using the Diva software and analyzed using FlowJo.

### Statistical analyses

#### Bulk transcriptome analysis

We quantified bulk RNA sequencing data using Salmon^79^ using default parameters and the human GRCh38 genome. To check for potential outliers, we performed principal component analysis (PCA). No samples were removed based on this analysis. We then imported the transcript-level quantifications and transcript lengths using tximport^80^ and quantified the differential expression between responder groups and over time points using a limma-voom approach^81, 82^: *gene* ∼ *responder + age + Time +sex + 1|donor. For time dynamics, following linear model was used: gene ∼ responderTime + age + sex + 1|donor.* Functional enrichment was performed using gene set enrichment analysis (GSEA) using blood transcription modules (BTMs)^23^. Genes were ranked by t-test statistic from this mixed linear model for the comparison of interest. P-values were adjusted using Benjamini-Hochberg, where we considered adj. p value < 0.01 significant or adj. p value < 0.05.

The relative contribution of individual cell types was estimated using CIBERSORT^24^. using the LMM22 background. We defined the pathway activity as the mean expression corrected for variance across donors of all genes assigned to a specific BTM per sample. A linear model was used to associate the estimated proportions of individual cell types to functional pathways: *cell proportion ∼ BTM_score + age + sex + 1|donor*, done separately for TRs and NRs. where *BTM_score* is the BTM score as described above and *cell proportion* is the estimated cellular proportions from CIBERSORT per individual. P-values were adjusted on all associations using Benhamini-Hochberg. We considered adj. p-values < 0.05 to be significant.

#### Targeted proteomics

We removed measurements that were flagged as unreliable by Olink as well as protein assays where the target protein was detected in less than 70% of the samples. Finally, we considered 311 high-quality proteome assays for further downstream analyses. We performed dimensionality reduction (PCA) to check for potential outliers or batch effects and did not identify either.

To quantify the differences in protein abundances between timepoints within each responder group separately, we fit a linear mixed model using limma^81^: *protein* ∼ *time_responder* + age + *sex* + *1|donor*, where *time_responder* indicates the sampling time point and responder group, and *donor* is the random effect. Benjamini-Hochberg post-hoc correction was used to control the false discovery rate across all proteins. We considered adjusted p-values < 0.05 significant.

#### Untargeted metabolomics

Raw metabolite profile data were centroided, merged and recalibrated using MATLAB software as previously described^76^. Putative annotations were generated based on compounds contained in the HMDB^83^ database using both accurate mass per charge and isotopic correlation patterns. We retained only ions that were confidently annotated, allowing 0.001 Da tolerance between the ion and its corresponding annotation. Since exogenous and/or drug-related metabolites do not reflect an individual’s current immune status and could lead to possibly confounding effects, we aimed to consider a set of endogenous metabolites. To do so, considered endogenous metabolites from HMDB and matched this to our annotated metabolite data based on the ion’s chemical formula. Then, we manually re-checked whether metabolites were drug-related using the DrugBank database^84^. Metabolites associated with medications or other xenobiotics were removed. Finally, we retained 192 endogenous metabolites for further analyses.

We validated the quality of our untargeted metabolome data by using a targeted metabolomic approach on a subset of the individuals. We manually correlated 18 primary amino acids that were reliably detected and annotated between the two platforms and observed good replicability (median Pearson’s *r* = 0.77). This indicates good sample quality and reliable, consistent measurements (Supplementary Figure 11).

We fit a linear mixed model in limma^81^ to quantify differences in endogenous metabolic abundances within responder groups over time: *metabolite* ∼ *time_responder* + age + *sex* + *1|donor*, where *time_responder* indicates the sampling time point and responder group, and *donor* is the random effect. We applied Benjamini-Hochberg post-hoc correction on all endogenous metabolites and considered metabolites with adjusted p-values < 0.05 significant. Metabolite classifications to annotate metabolites were retrieved from HMDB. We performed metabolite enrichment by performing over-representation analysis using IMPALA^85^. Adjusted p-values (Benjamini-Hochberg) < 0.05 were considered significant.

#### Integration of transcriptome, proteome and metabolome layers

We aimed to provide a comprehensive integration of the different data layers that were generated. To do so, we employed two approaches. First, we associated circulating protein and metabolite levels to transcriptome pathway activity. We defined pathway activity as described above. Proteins and metabolites were selected when differentially abundant at Day 7 compared to Day 0. Associations were estimated using a linear mixed model: *protein ∼ age + gender + BTM + 1|donor.* P-values were adjusted using Benhamini-Hochberg over all obtained associations, and adjusted p-values < 0.05 were considered significant. We performed this analysis for Day 7 as we observe the peak of the adaptive immune response to vaccination on this day.

Next, we used Multi-Omics Factor Analysis (MOFA)^37^ to perform unsupervised integration of all layers. We chose day 0, day 7 and day 35 as we had datasets for these time points across all layers for the highest (N = 10) and lowest (N = 10) responders. For transcriptome, the top highly variable genes were selected. For proteome and metabolome, we removed molecules that were significantly (nominal p-value < 0.05) associated with sex. Finally, we considered 1560 genes, 279 proteins and 158 metabolites across three timepoints for further analysis using MOFA. We applied MOFA standard practices and trained the model on a single group. The default data and training model options were used except for scale_views set to TRUE. Following model training, MOFA reported factor 1 correlated with technical variation and we thus did not consider this factor. The rest of the factors were evaluated for significance with either Responder Category, Age, Gender and Time by testing the factor values using a Wilcoxon ranked-sum test. For the factors that are shown, weights are scaled for each layer independently and used to identify the top predictors for proteome and metabolome. For network building, we performed GSEA using BTMs on the transcripts ranked by MOFA’s scaled weights, and considered adjusted p-values < 0.05 significant. For proteome and metabolome, molecules with absolute scaled weights > 0.25 for factor 3 were considered and their associations done, similar to BTMs. Associations across layers of protein and metabolite molecules to different BTMs controlling for repeated measurements from the same donors were done and associations with adjusted p-values < 0.05 were selected. Across-layers associations were also calculated and significant associations were kept. Cytoscape^86^ was used to build the multi-layer network with colour of the nodes representing factor 3 scaled weights for each protein or metabolite, or enrichment score for transcriptome BTMs.

#### Partial least-squares regression

We employed partial least-squares regression (PLSR) to identify the pre-vaccination proteins and metabolites that are most consistently associated with the serological response against the three influenza strains. PLSR was performed using the *pls* library in R for proteome and metabolome separately using all 200 donors of the discovery cohort. The model was run for 100 independent iterations with 10-fold cross-validations (e.g. 90% training, 10% testing) and we calculated the proportion of variation explained by each PLSR component. For the first 3 components we performed rank-product analysis over the 100 iterations to identify the top predictors. The top 30 ranked molecules (either proteins or metabolites) were then evaluated for their positive or negative association with antibody fold change.

## SUPPLEMENTAL INFORMATION TITLES AND LEGEND

**Supplementary Figure 1** *General demographic factors associated to the serological response to trivalent inactivated influenza vaccination.* **(A)** Correlation among hemagglutination (HAI) titers fold-change upon vaccination. In both seasons, HAI titers against each of the three strains are moderately positively correlated to each other. **(B)** Correlation between MN titers and HAI titers across two influenza seasons. Each dot is a sample, colours represent influenza strains. **(C)** Impact of sex on the serological response to TIV across two seasons. P-values were generated using the Wilcoxon ranked-sum test. **(D)** Influence of age on the HAI and microneutralization (MN) titers. Each dot is an individual, colours represent the different influenza strains.

**Supplementary Figure 2** *Differences in cellular populations between responders and non-responders.* **(A)** Deconvoluted cellular populations were estimated using CIBERSORT from bulk transcriptomics data (n=10 TR, n=10 NR) across five different timepoints. Each point indicates a sample (NR=blue, TR-red). The curve is a smooth fitted line with standard error of the estimation. **(B)** CBC at T1 (pre-vaccination) for Neutrophils, Leucocytes and Monocytes for 200 donors. Samples are stratified by their responder status: TR, Other or NR.

**Supplementary Figure 3** *Time dynamics and BTMs in TRs and NRs.* **(A)** Significant BTMs upregulated in TRs and NRs at pre-vaccination stage. **(B)** Genes enriched for inflammatory BTMs and their expression in TRs and NRs. Only CD83 and IL23A are significantly (adjusted p-value < 0.05) upregulated in TRs **(C)** Number of significantly differentially expressed genes in TRs and NRs, 3days, 7 days, 35 days and 70 days post vaccination **(D)** Top 20 significant BTMs upregulated and downregulated in TRs 3 days post vaccination. **(E)** BTMs significantly upregulated in TRs (left), NRs (right) 7 days post vaccination.

**Supplementary Figure 4** *NK cell flow cytometry results from the replication cohort.* Total NK cell proportions (top), CD56dim (middle) and CD56bright (bottom) flow cytometry results from NRs (n=2, except day 6/7 where n=1) and TRs (n=5).

**Supplementary Figure 5** *Proteomic and metabolomic response to TIV.* **(A)** CLECL7A proteome abundance in all TRs (red) and NRs (blue) over time in the Discovery cohort (top) and the Replication cohort (bottom). **(B)** Proteome differential abundance results in the replication cohort. **(C)** Volcano plots showing differential abundance results comparing Day 35 vs Day 0 for TRs (left) and NRs (right) in discovery cohort. Red dots represent significantly different proteins at adjusted p-value < 0.05. **(D)** Annotation of the differentially abundant metabolites found in the replication cohort. Baseline represents the universe of all metabolites measured, whereas TR and NR show significantly differentially abundant metabolites (nominal p < 0.05) at T3 and T4, respectively. (**E)** Subclass annotation of upregulated organic acids (top) and lipids and lipid-like molecules (bottom). These metabolites were significantly differentially abundant (adjusted p < 0.05) in the discovery cohort. Pie charts indicate the distribution of significant metabolites within the fatty acids and conjugates classification. **(F)** Alluvial plot showing the differential expression results for TRs (left) and NRs (right) at the different timepoints compared to baseline in the discovery cohort (n=81). Significant metabolites shown (adjusted p-value < 0.05).

**Supplementary Figure 6** Proteins upregulated in NRs 7 days post vaccination and their correlation with transcriptome chronic inflammatory BTMs. Most proteins show a negative correlation with unadjusted p-value < 0.05.

**Supplementary Figure 7** *Multiomics integration capturing different dynamics in TRs and NRs.* **(A)** Factor 4 and Factor 6 showing time variation captured in TRs. Factor 4 is exclusively explained by transcriptome while factor 6 is explained by both transcriptome and proteome. **(B)** Abundance of top factor 3 proteins and metabolites in all 81 TRs and NRs. **(C)** Methionine positive correlation with cytokine production post influenza stimulation in 500FG cohort. **(D)** Distribution of scaled weights attributed to proteins, metabolites and enrichment score for transcripts. Molecules with scaled weights of absolute value > = 0.25 were considered for network generation in Figure 4E.

**Supplementary Figure 8** *Pre-vaccination correlates* (**A)** Plot of TNFSF13/APRIL against log2 antibody fold change of each strain in discovery (left) and replication cohort (right). **(B)** Plot of IL15 against antibody fold change of each strain in replication cohort. **(C)** Expression of IL-15 in human PBMCs using single-cell RNA sequencing data.

**Supplementary Figure 9** *IL15RA-/-mice experiments* **(A)** Representative flow cytometry plots showing the frequency of B cells (CD19+) and T cells (CD3+) in the spleen of IL15RA^−/−^ mice (left plot), and the NK cell frequency (NK1.1+) among splenocytes from IL15RA^+/+^ and IL15RA^−/−^ mice (two plots on the right). **(B)** The dot plots represent the frequency of total B cells, GC B cells, total CD4+T cells, CD4+Foxp3-Tconv cells, CD4+Foxp3+ Treg cells, Tfh, and Tfr cells in inguinal lymph nodes from IL15RA+/+ and IL15RA-/-mice. Pooled data from two independent experiments; n=8 to 10.

**Supplementary Figure 10** *Pre-vaccination metabolite correlates with antibody fold change.* **(A)** Plot of citric acid against antibody fold change for each strain. **(B)** Plot of citric acid abundance against cell proportions calculated from 300BCG cohort. **(C)** Plot of betaine(top) and cysteine(bottom) abundance against antibody fold change for each strain. **(D)** Additional unsaturated long chain fatty acids negatively correlated to IL15. **(E)** Estimate of protein (42) associations to all fatty acids for ∼200 elderly donors (left) and to ∼ 500 young donors (right). In the elderly, LCFA negatively correlated to most proteins while other fatty acids (FAs) show both positive and negative correlation while no such pattern observed in the younger cohort. Associations with p-value <0.05 plotted.

**Supplementary Figure 11** *Replicability of untargeted metabolomic profiles.* We manually correlated 19 primary amino acids that were confidently detected in both the untargeted and targeted metabolomics dataset. Each dot indicates a sample, coloured for time (10TRs, 10NRs across three timepoints). We observe good replicability for the primary amino acids.

