## Supplemental Figures for "Systems immunology reveals the molecular mechanisms of heterogeneous influenza vaccine response in the elderly"

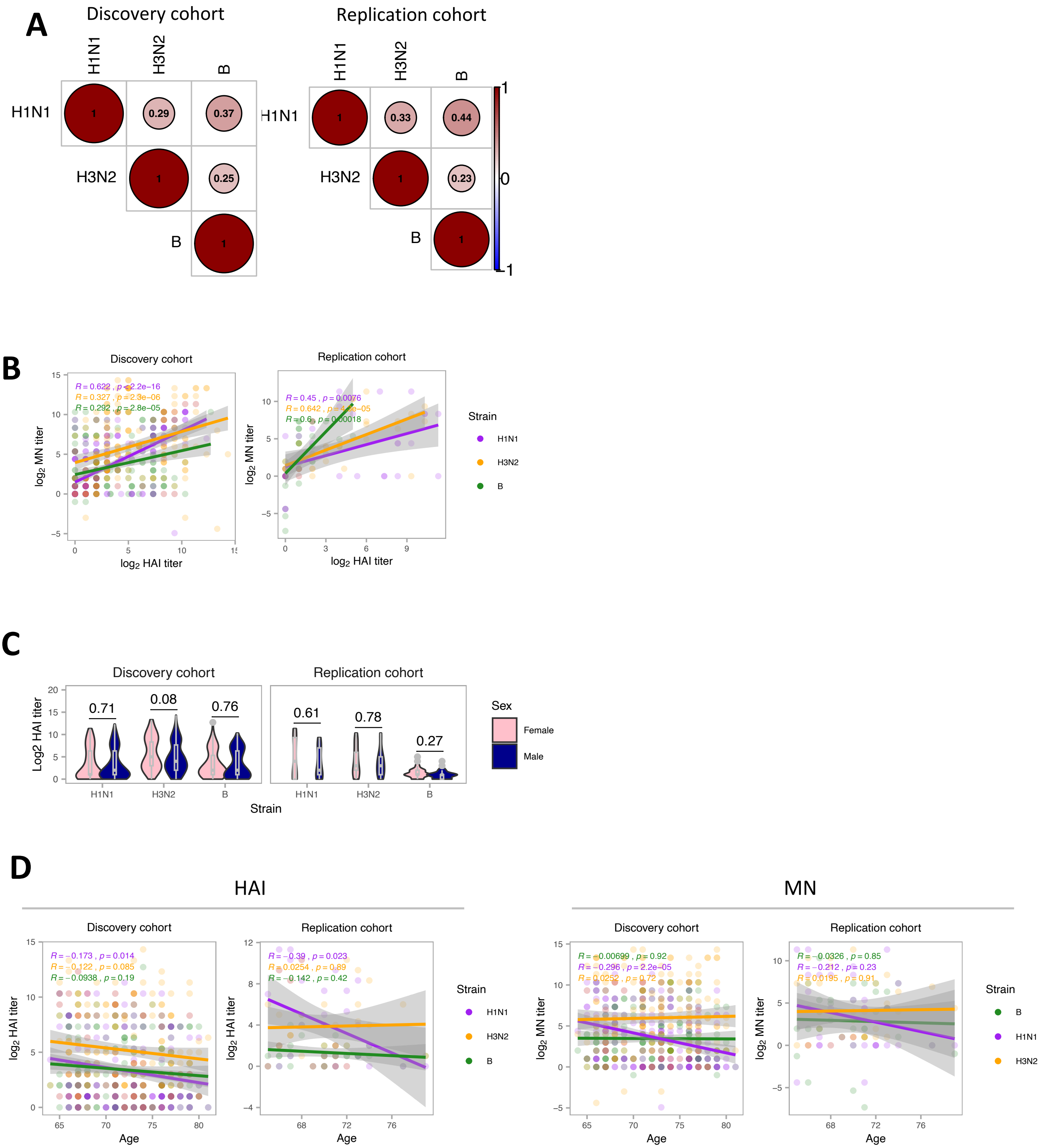


Supplementary Figure 1

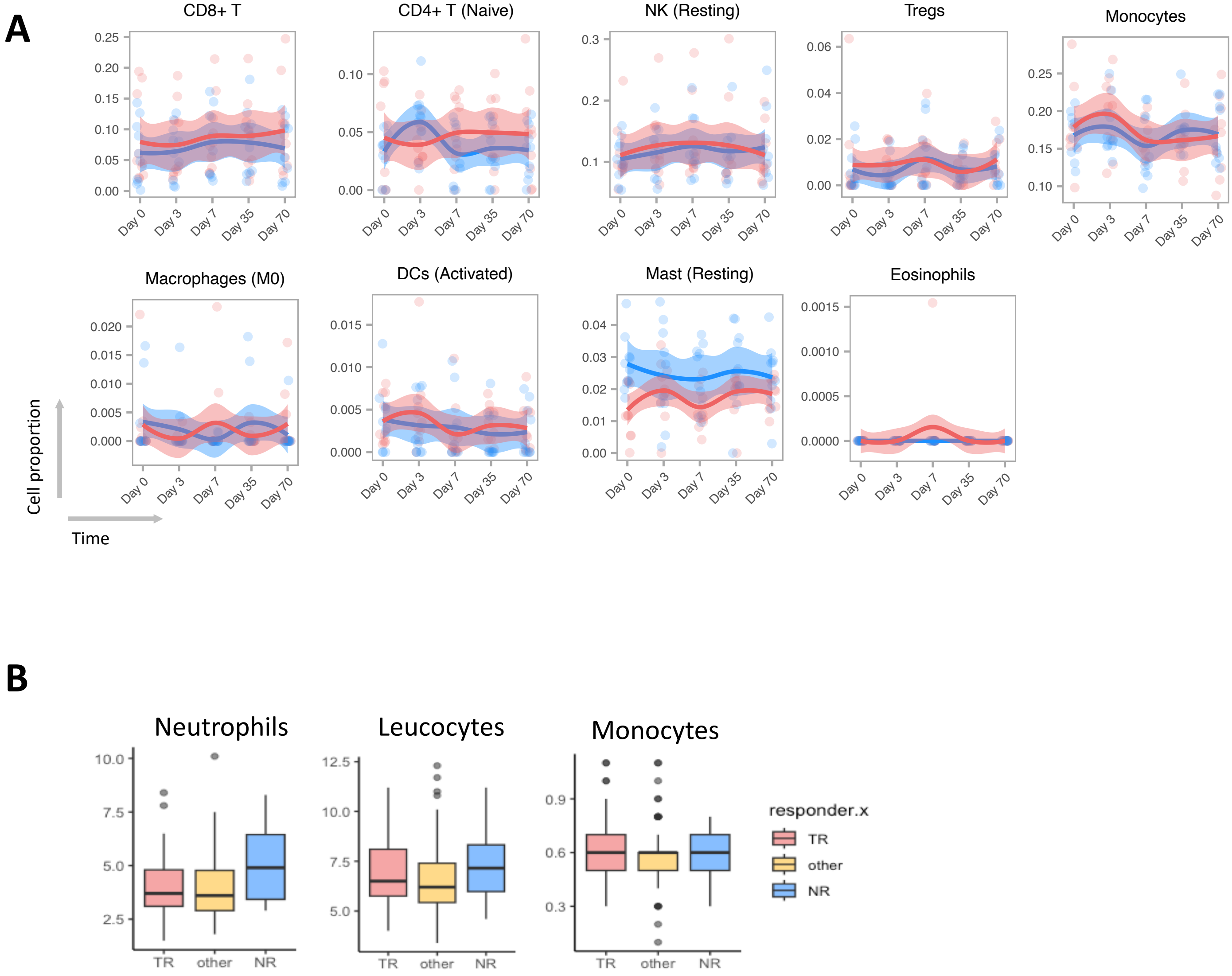


Supplementary Figure 2

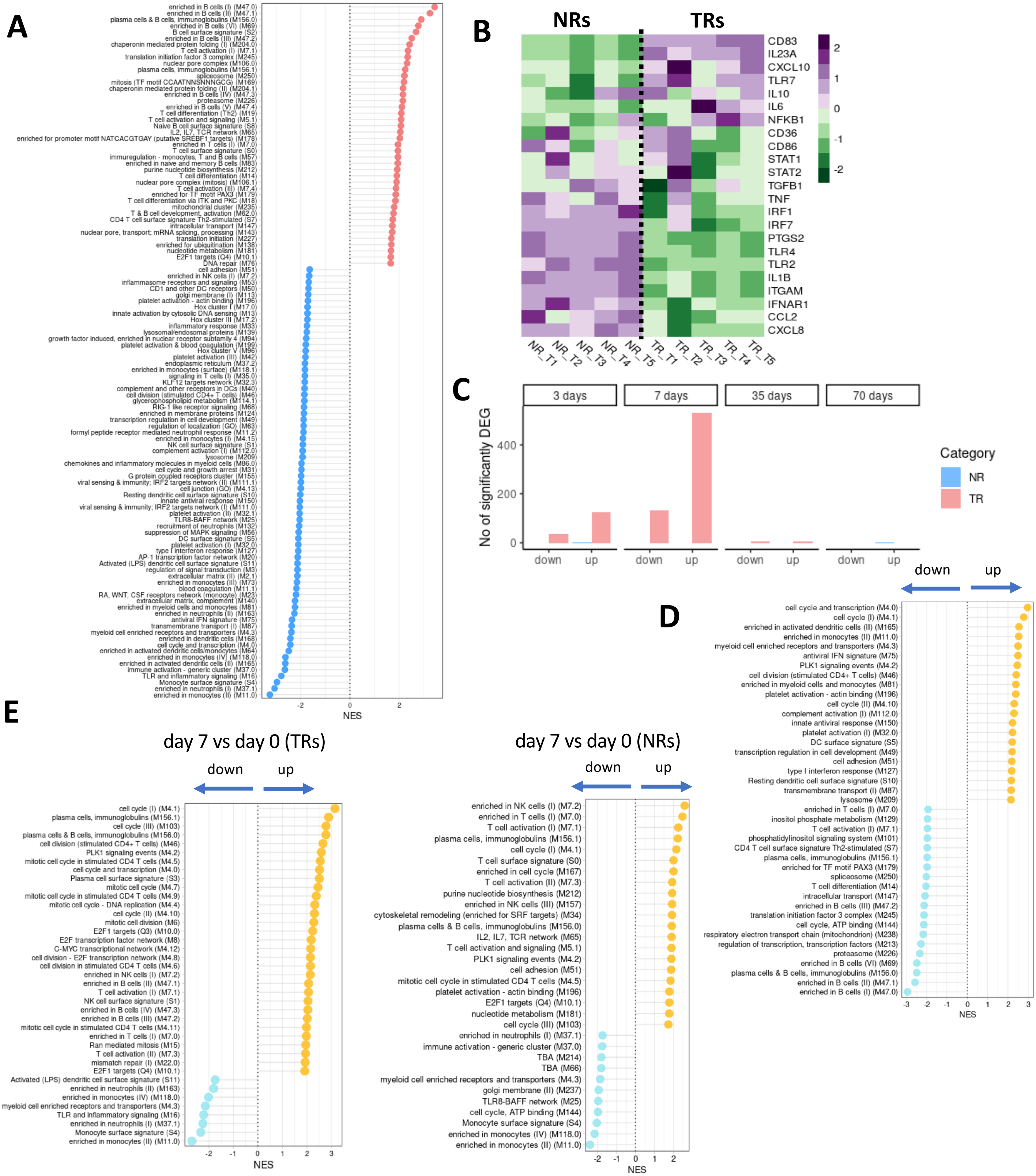


Supplementary Figure 3

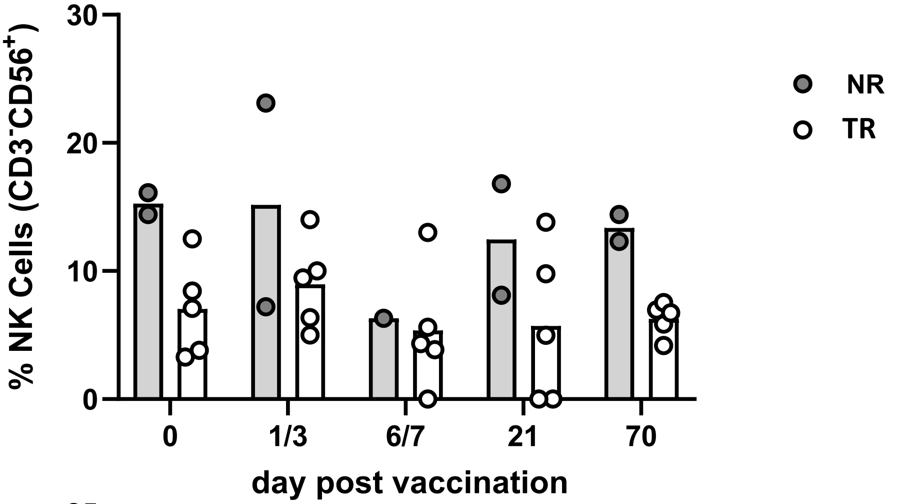


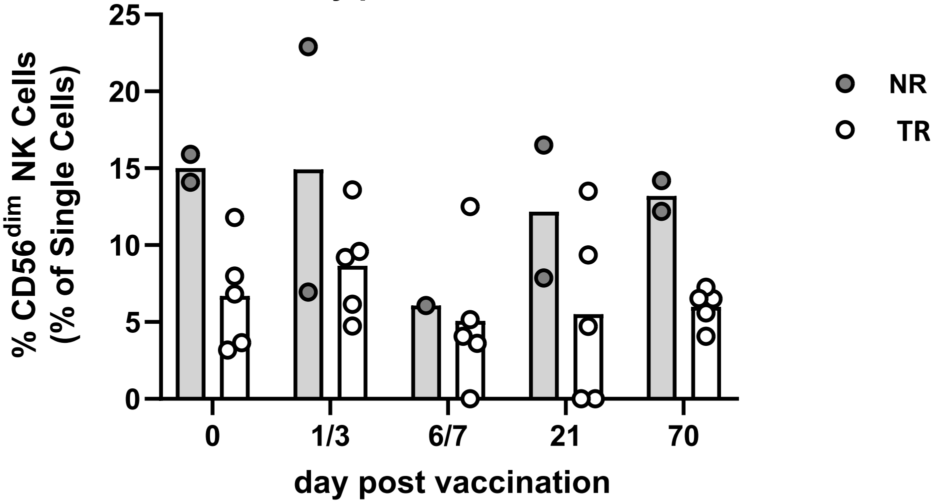


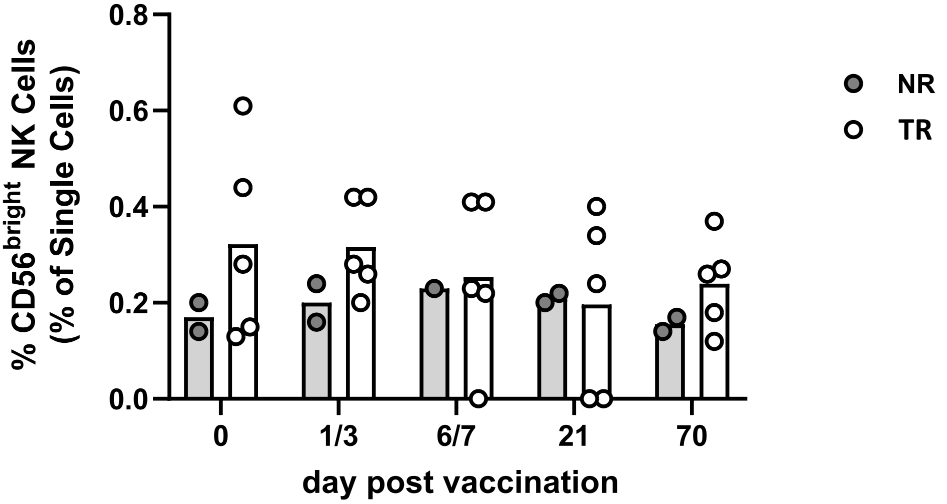


Supplementary Figure 4

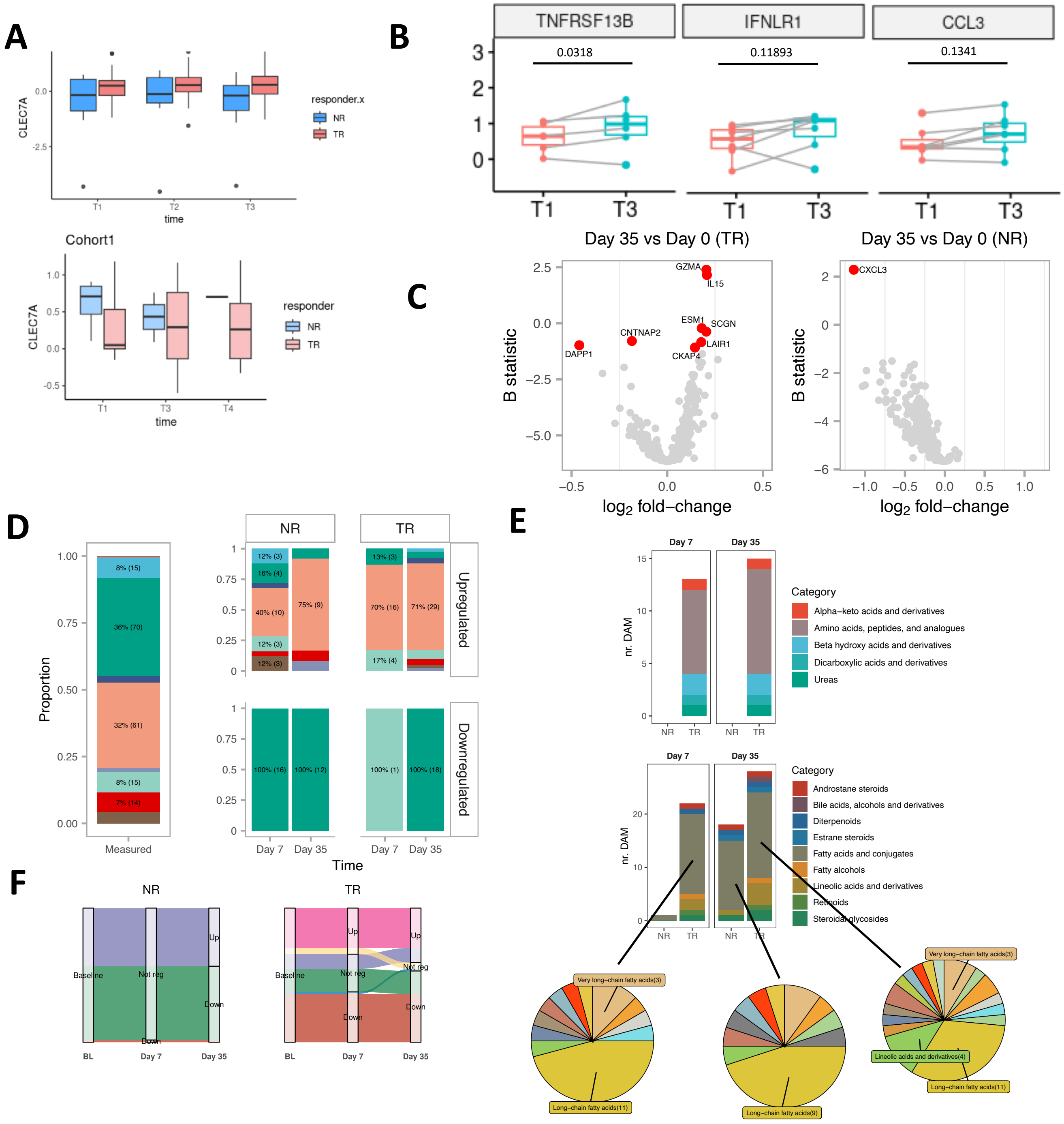


Supplementary Figure 5

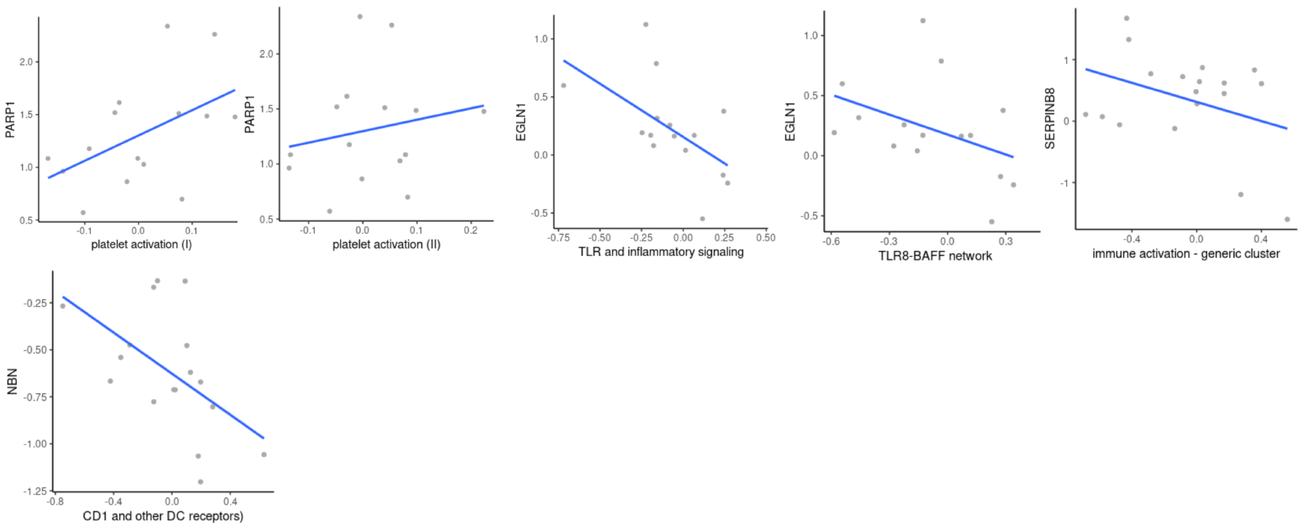


Supplementary Figure 6

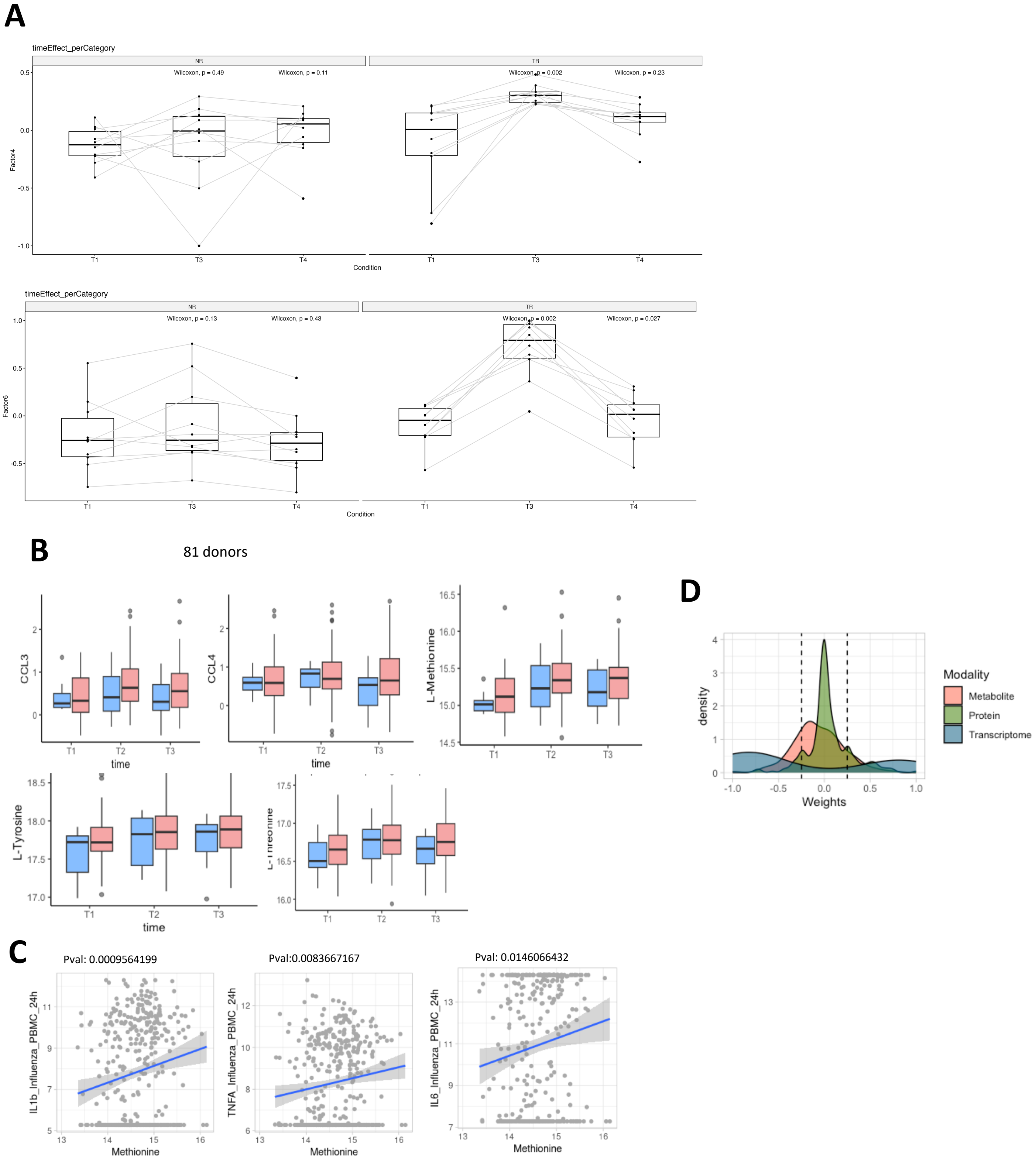


Supplementary Figure 7

A

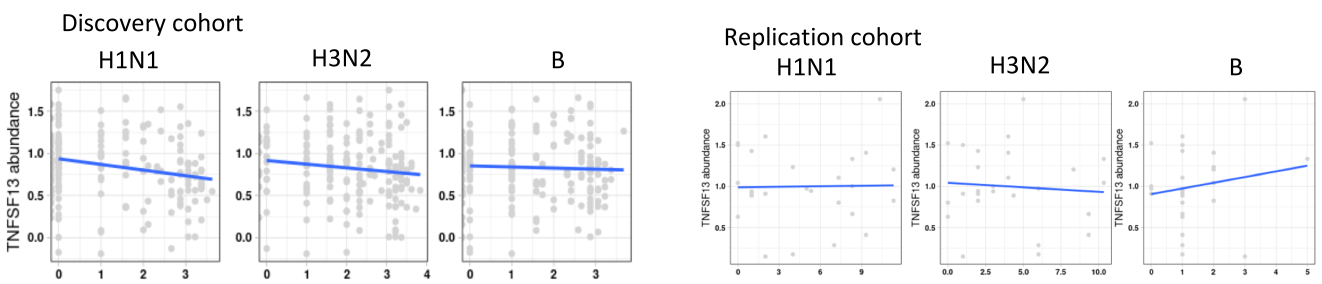


B

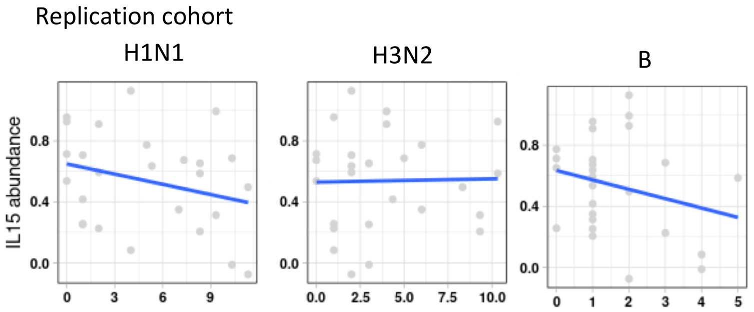


C

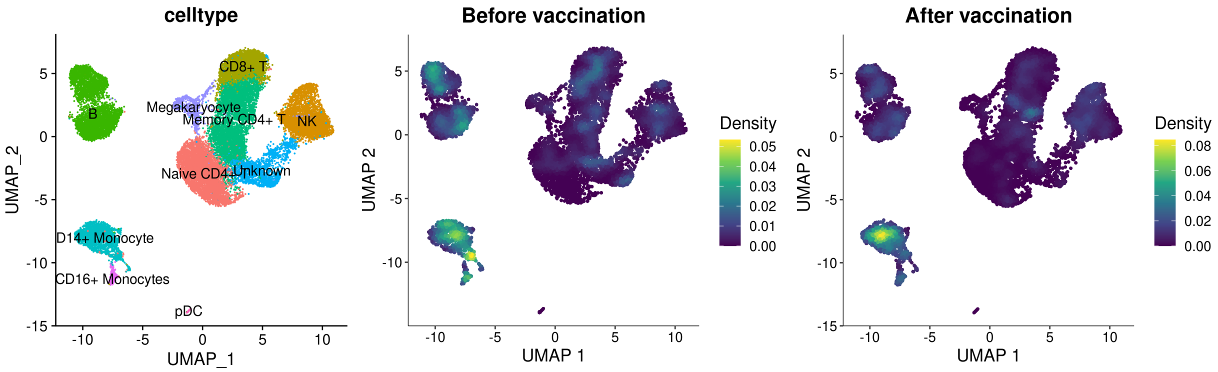


Supplementary Figure 8

A

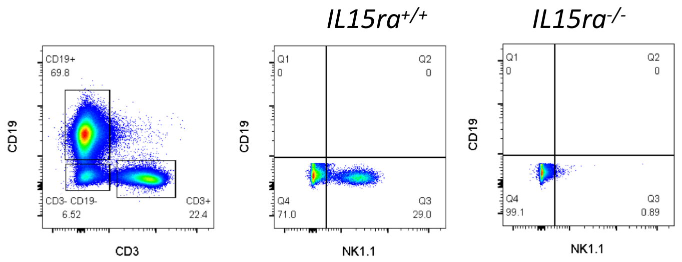


B

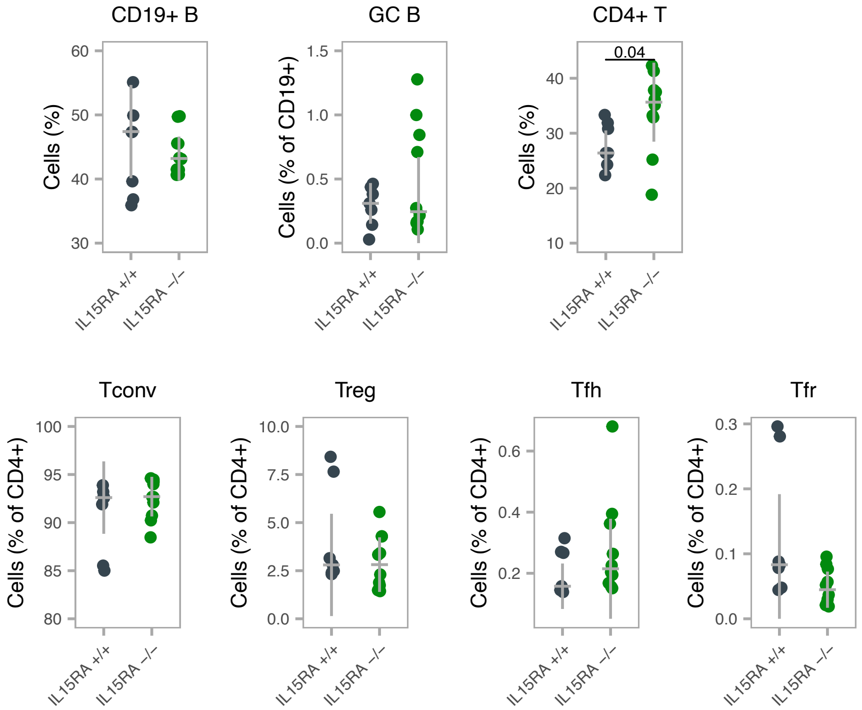


Supplementary Figure 9

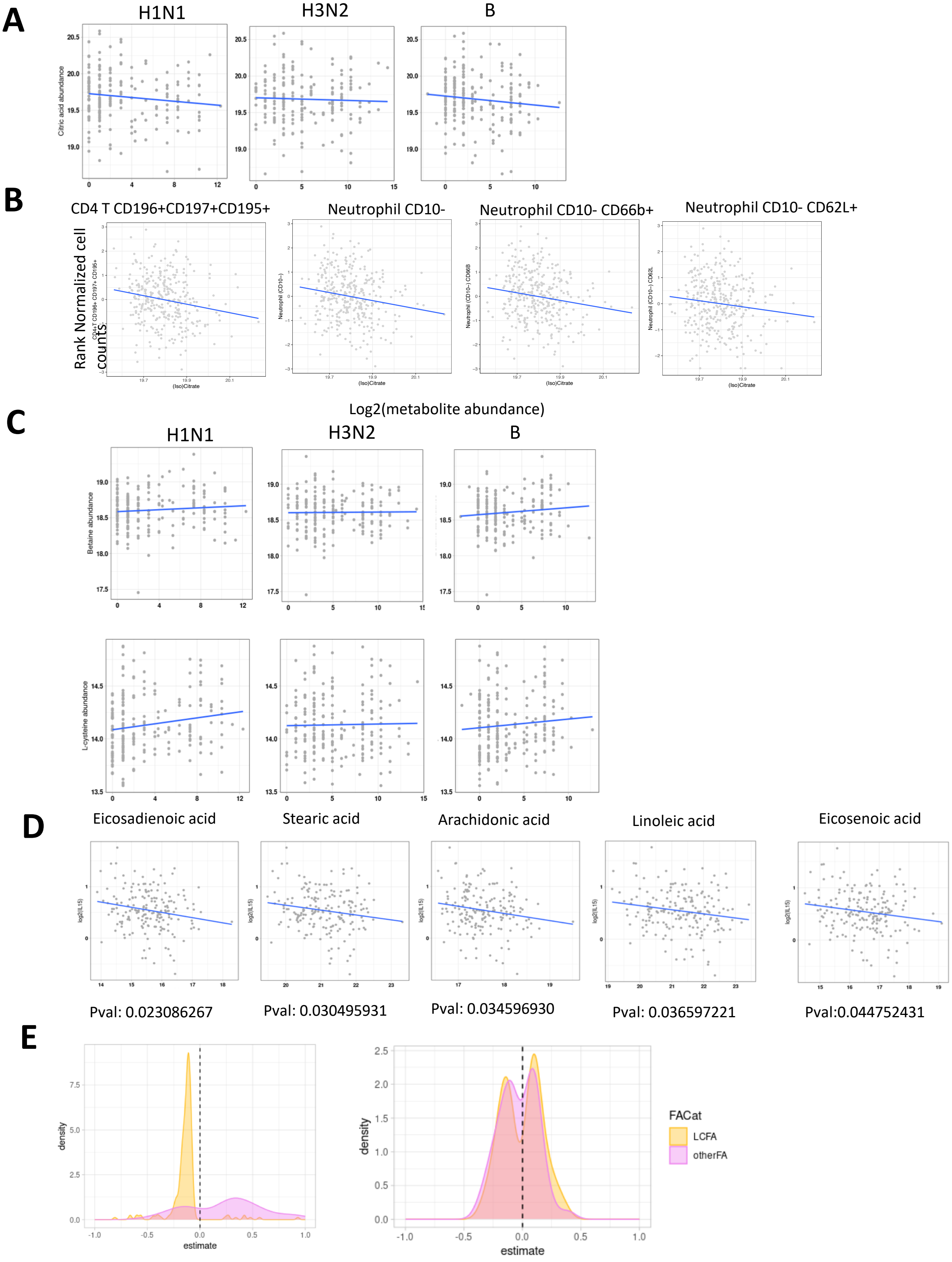


Supplementary Figure 10

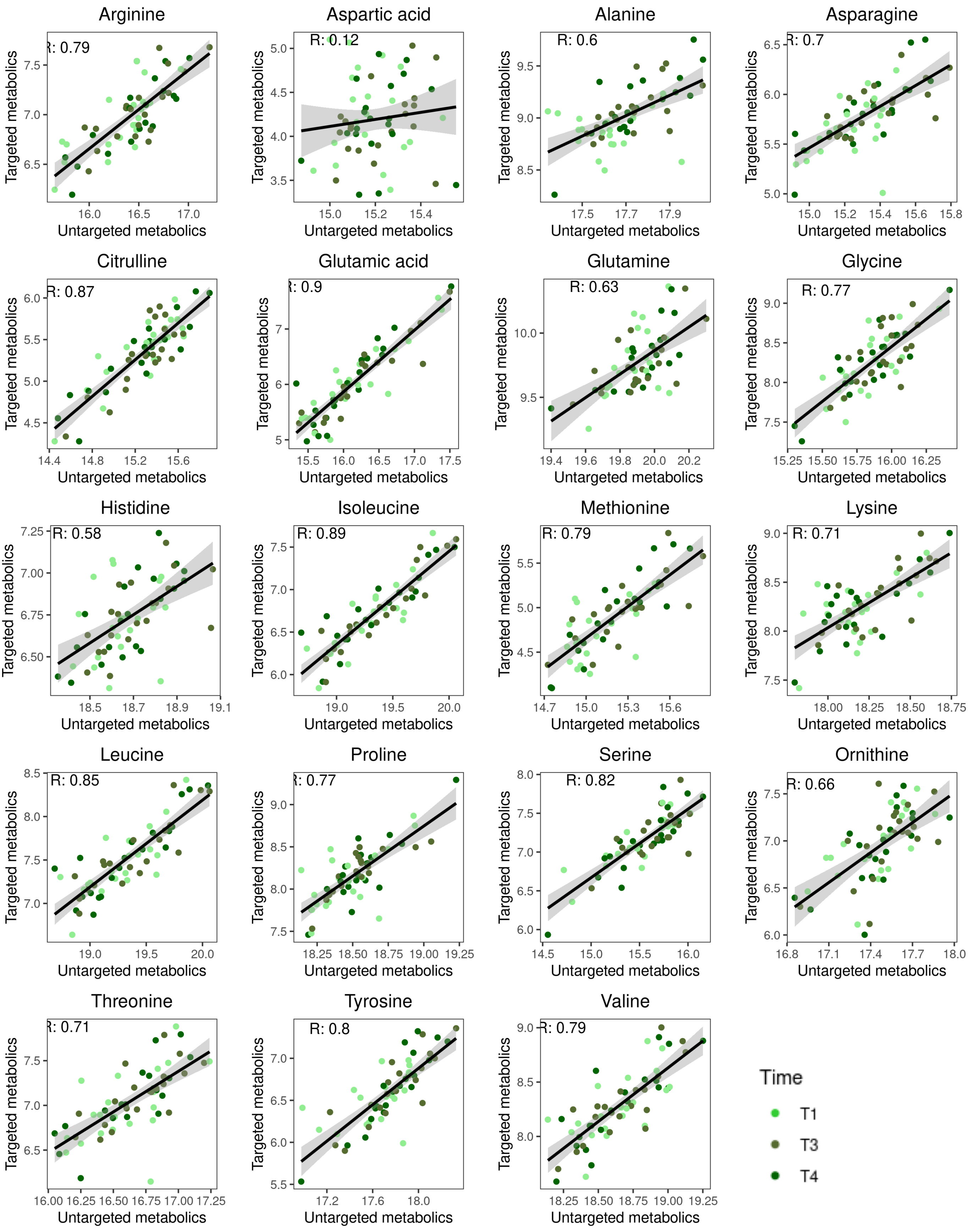


Supplementary Figure 11
